# Dissecting Biological Pathways of Psychopathology using Cognitive Genomics

**DOI:** 10.1101/2021.07.20.21260487

**Authors:** Max Lam, Chia-Yen Chen, W. David Hill, Charley Xia, Ruoyu Tian, Daniel F. Levey, Joel Gelernter, Murray B. Stein, Biogen Biobank team, Alexander S. Hatoum, Hailiang Huang, Anil K. Malhotra, Heiko Runz, Tian Ge, Todd Lencz

## Abstract

Cognitive deficits are known to be related to most forms of psychopathology. Here, we perform local genetic correlation analysis as a means of identifying independent segments of the genome that show biologically interpretable pleiotropic associations between cognitive dimensions and psychopathology. We identified collective segments of the genome, which we call “meta-loci”, that showed differential pleiotropic patterns for psychopathology relative to either *General Cognitive Ability* (GCA) or *Non-Cognitive Skills* (NCS). We observed that neurodevelopmental gene sets expressed during the prenatal-early childhood predominated in GCA-relevant meta-loci, while post-natal synaptic gene sets were more involved in NCS-relevant meta-loci. Notably, we found that GABA-ergic, cholinergic, and glutamatergic genes drove pleiotropic relationships within dissociable NCS meta-loci.

---

Cognitive impairment is one of the core features of psychopathology and is associated with the debilitating nature of many psychiatric disorders^1,2^ In schizophrenia, for example, cognitive impairments are predictive of known functional impairments even in the prodromal stage of the illness^3,4^. Cognitive deficits are not only confined to adult psychiatric illnesses, but also extend to childhood disorders like Autism Spectrum Disorders (ASD) and Attention-Deficit Hyperactivity Disorder (ADHD)^5^. Individuals who suffer from psychiatric disorders tend to report sequelae of cognitive problems throughout their lifetime^6^. In many cases, there is an emergence of cognitive deficits before a formal diagnosis of mental illness^7^.

Prior to the era of well-powered GWAS in psychiatry, the idea of using cognitive function as an endophenotype to understand the biology of psychopathology was proposed^8^. We presented the first molecular genetic evidence for overlap between general cognitive ability and schizophrenia^9^. Since then, evidence suggesting widespread pleiotropy across psychopathologic traits has emerged, indicating shared biological mechanisms^10,11^. Pleiotropy, a phenomenon where a genetic variant might affect several traits at once, appears to be ubiquitous in biology; 44% of the loci reported within the GWAS catalog have been shown to be associated with more than one trait^12^ (although in some cases this may be a function of linkage disequilibrium rather than true pleiotropy^13^). A more recent study indicated that trait associated loci cover more than half of the genome, among which 90% implicate multiple traits^14^.

We recently exploited pleiotropy to dissect biology underlying the counterintuitive positive genetic correlation between educational attainment and schizophrenia, and were able to parse separate neurodevelopmental and synaptic mechanisms underlying the disorder, based on association patterns within GWAS significant loci^15^. Shortly after, Demange and colleagues^16^ demonstrated that it was possible to leverage global genetic correlation within a structural equation modelling framework to derive a latent *Non-Cognitive Skills* (NCS) factor by removing variance related to *General Cognitive Ability* (GCA) from educational attainment GWAS. The ensuing NCS factor showed positive genetic correlation with schizophrenia, consistent with our earlier findings. These two reports, utilizing somewhat orthogonal but complementary approaches, leveraged pleiotropic phenomena to study the intersection of cognition and psychopathology. Nevertheless, these studies are limited by following a global genetic correlation approach^17^ on the one hand, and a SNP-by-SNP approach^15,16^ on the other. As has been demonstrated^18^, the assumption that genetic correlations for complex traits are homogeneously distributed across independent genomic regions may not be true. At the same time, typical locus-based GWAS comparisons tend to be defined by “top” SNPs followed by LD clumping; this invariably results in loss of information from regions of the genome that fall short of genome-wide significance. Studies using partitioned heritability or gene set analysis have demonstrated the biological relevance of regions of the genome that do not contain genome-wide significant loci^19^. In the present study, we develop a novel method, intermediate to global and SNP-based approaches, that examines the structure of local genetic correlations across the genome and identifies “meta-loci”, which we define as combined genomic segments sharing pleiotropic patterns. We then interrogate these meta-loci, using rich annotations and gene set analysis, to identify and dissociate biological pathways underlying different patterns of cognitive-psychopathologic pleiotropy.

## Results

### Study Design and Methods Overview

We have recently reported the largest GWAS meta-analysis for GCA (N=373,617) and utilized this well-powered phenotype for pleiotropic analysis with an expanded list of psychopathology phenotypes^20^. Expanding on earlier analytic strategies^15,16^, we carried out three broad stages of analyses (Fig-1).

**Figure 1.**
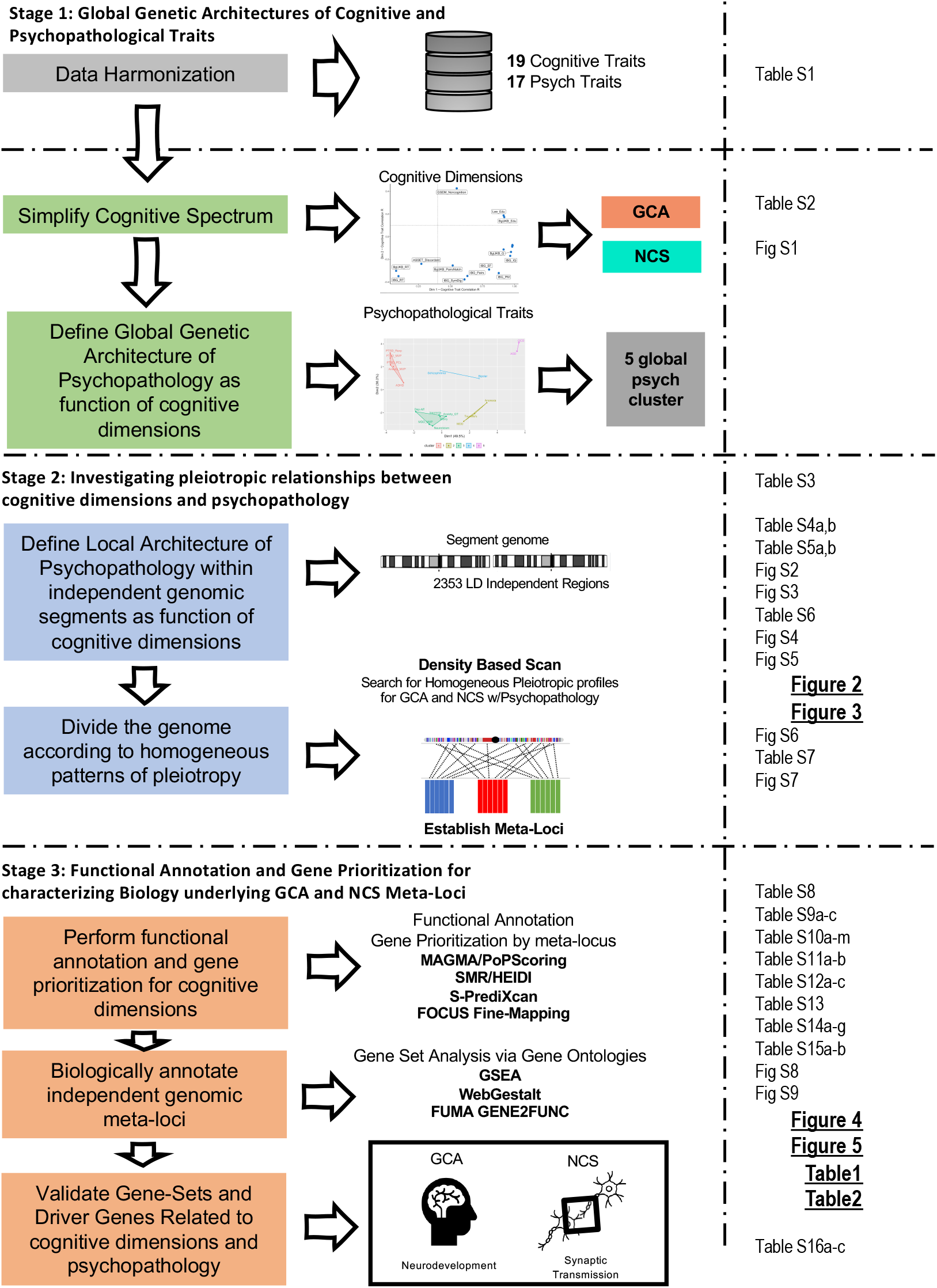
Study Procedures and Analytic Workflow. ***Note:*** The flow chart is divided in three columns. The first column from the left describes the analytic objectives across study stages. The middle column lists the method/software used for the analysis. And the right column points the reader to the Table, Figure, Supplementary Table or Supplementary Figure that reports the results of the analysis.

First, as both a data reduction step and a benchmark for subsequent steps, we examined the global pleiotropic relationships between multiple cognitive and psychopathological phenotypes via LD score regression^21,22^. We amalgamated all available summary statistics from recent GWAS of cognitive function (N = 19). These included GCA^20^, and other reported derivatives of cognition including NCS^16^, Executive Function^23^, and GWAS of individual cognitive tests administered as part of the UK Biobank. Similarly, traits related to psychiatric illnesses or psychopathology in the recent studies from the Psychiatric Genomics Consortium, UK Biobank, Million Veterans Program and elsewhere were curated (N = 17; Table-S1). Data reduction approaches indicated that it was appropriate to focus on GCA and NCS traits for the purpose of the current study (Fig-S1). We re-estimated NCS using the largest GCA GWAS^20^ to increase statistical power (Supplementary Information).

Second, we carried out local genetic correlation analyses between each of the psychopathology traits and GCA, and separately between each psychopathology trait and NCS. A series of positive and negative local genetic correlation patterns emerged across these analyses, which we then classified via the “meta-locus” approach. A meta-locus was defined as a set of LD-independent regions that showed similar local genetic correlation profiles across psychopathological traits across the genome. There were 15 distinct meta-loci (Median Length = 34.24 Mb) identified for GCA and 12 identified for NCS (Median Length = 23.19 Mb).

Third, GCA and NCS GWAS summary statistics were functionally prioritized to identify genes and biological mechanisms harbored within the meta-loci. A series of Gene-Based Genome-Wide Association (GBGWA) and Transcriptome-Wide Association (TWA) strategies were applied to GCA and NCS GWAS summary statistics. We leveraged brain eQTLs from a range of databases, including GTEx v8 brain tissue expression^24^, Brain-eQTL-meta^25^, and PsychEncode^26,27^ eQTL databases that indexes brain (Online Methodology and Supplementary Information). We adopted a broadly inclusive approach to the gene prioritization stage of the analysis, excluding from consideration only those genes with no supporting evidence from any of these procedures. Next, we performed a series of gene set analyses^28–31^ and gene scoring procedures^32^, to which we applied strict filtering criteria in order to arrive at a high-confidence biological characterization of each meta-locus. We evaluated the high-confidence genes emanating from the GBGWA, TWA and gene set analysis approaches for longitudinal gene expression across lifespan to further differentiate neurodevelopmental vs adult synaptic mechanisms. Finally, we annotated driver genes with information on the propensity for psychiatric or nootropic drug re-purposing (Fig-1, Online Methodology).

#### Stage 1: Global genetic architectures of cognitive and psychopathological traits

A wrapper function within GenomicSEM^11^ was used to conduct LD score regression^21,22^ and create a global 19 × 17 genetic correlation matrix of cognitive and psychopathological traits (Table-S2). Two-dimension reduction techniques, PCA and partitioned cluster analyses (K-Medoid) dimension reduction techniques were applied to the global genetic correlation matrix to identify underlying association patterns. The PCA resulted in a “Cognitive Map” (Fig-S1a). The first principal-component captured the similarity of each cognitive trait to general cognitive ability (GCA) in context of its relationship to the vector of 19 psychopathological traits (Fig-S1b). The second principal-component represented the degree to which a cognitive trait is similar to NCS given its relationship to the vector of 19 psychopathological traits (Fig-S1c). Consequently, for all subsequent analyses, we focused on GCA and NCS as the primary cognitive phenotypes of interest; we leave a more detailed exploration of the cognitive phenotypic space to future work. We re-extracted the NCS latent factor by using GWAS-by-subtraction parameters^16^, and a better powered GCA meta-analysis^20^. We confirmed that our newly calculated GCA and NCS factors were globally similar to those originally reported by Demange et al.^16^ (r_g_=1 using LD score regression). We utilized the current versions of GCA and NCS in subsequent sections detailing functional annotation, gene prioritization and gene set analyses. At the global level, we also observed that psychopathological traits separated into five best fitting clusters (Table-S2c) that varied according to the degree of the strength of relationship with GCA and NCS (Fig-S1d, inset). There was much more variation across psychopathological traits than the cognitive traits, so we decided to analyze each trait separately in the local genetic correlation analyses described below. However, we utilized the clustering results depicted in Fig-S1d as background information for our interpretations of subsequent results (and as color-coding in subsequent figures in this report).

#### Stage 2: Investigating pleiotropic relationships between cognitive dimensions and psychopathology

Local genetic correlation analyses were carried out across 2353 LD-independent regions of the genome using ρ-HESS^18^. 4,469,149 SNPs were included for GCA and 4,372,398 SNPs were included for NCS. Region specific heritability estimates of GCA and NCS were expectedly small (median *h*^*2*^_GCA-Region_ = 8.36e-5, median *h*^*2*^_NCS-Region_ = 1.18e-4, Table-S3). The sum of heritability across LD independent regions was consistent with previous reports for these phenotypes (*h*^*2*^_GCA_ *=* 0.23, *h*^*2*^_NCS_ = 0.31) (Supplementary Information).

We observed widespread pleiotropy for both GCA and NCS with psychopathology (Table-S4/S5). In general, global estimates of genetic correlations (via LD-score regression) were highly comparable to summed local estimates derived from ρ-HESS, except in the case of Anorexia Nervosa and OCD, for which ρ-HESS recovered a greater degree of correlation with NCS (Fig-S2). Aside from re-capitulating global R_g_ trends across psychopathology phenotypes, several noteworthy observations emerge from the local genetic correlations. First, for each psychopathological trait, local genetic correlations were not in the expected direction, across LD-independent regions, indicated by global genetic correlations (Fig-S3a/b). Second, the magnitude of the local genetic correlation signals is markedly stronger for schizophrenia than for any other trait. Third, schizophrenia is the only trait (potential exception of Anxiety_MVP) that demonstrates strong, widespread differences in direction between GCA and NCS correlations (Fig-S2, Table-S6). Fourth, local genetic correlations for GCA were especially strong within the Major Histocompatibility Complex (MHC) (See Fig-S4). Notably, anorexia nervosa and ADHD show the opposite pattern of local genetic correlations within the locus relative to other psychopathological traits.

Local genetic correlation profiles were further parsed to better understand pleiotropic patterns across cognitive function and psychopathology. UMAP dimension reduction, followed by clustering (using density-based scanning, DBscan) was applied to the matrix of local genetic correlations (17 psychopathologic traits x 2330 LD-independent regions, excluding the MHC) to identify clusters of LD-independent regions with distinct pleiotropic patterns for GCA or NCS across the various psychopathologic traits (see Online Methodology). Resulting clusters of LD-independent regions were termed “Meta-Loci”. We identified 15 meta-loci for GCA and 12 meta-loci for NCS (Fig-2a/b; see Supplementary Information for further details of the DBscan method). Critically, the extracted meta-loci were not simply a function of highly localized effects but were distributed across the genome (Fig-2c/d). Each meta-locus encompassed between 7 - 1125 (GCA; median = 28) and 8 - 1926 (NCS, median=16) LD-independent segments of the genome (total length 6.96 - 1340.79 Mb, median=34.24 Mb for GCA; 8.70 - 2290.00 Mb, median=23.19 Mb for NCS). For both GCA and NCS, the first meta-locus extracted (GCA-1 and NCS-1, respectively, indicated in grey in Fig-2), consists of all genomic segments that could not be further clustered by the DBscan search without being listed as outliers, such that the entire genome is accounted for. We prioritized meta-loci based on their relative contribution to their total heritability: 8 meta-loci for GCA explained 95% of the total heritability of GCA (GCA-1,3,4,5,6,7,8,12); similarly, 4 meta-loci for NCS were prioritized (NCS-1,2,3,5) (Fig-S3, Table-S4b/S5b).

**Figure 2.**
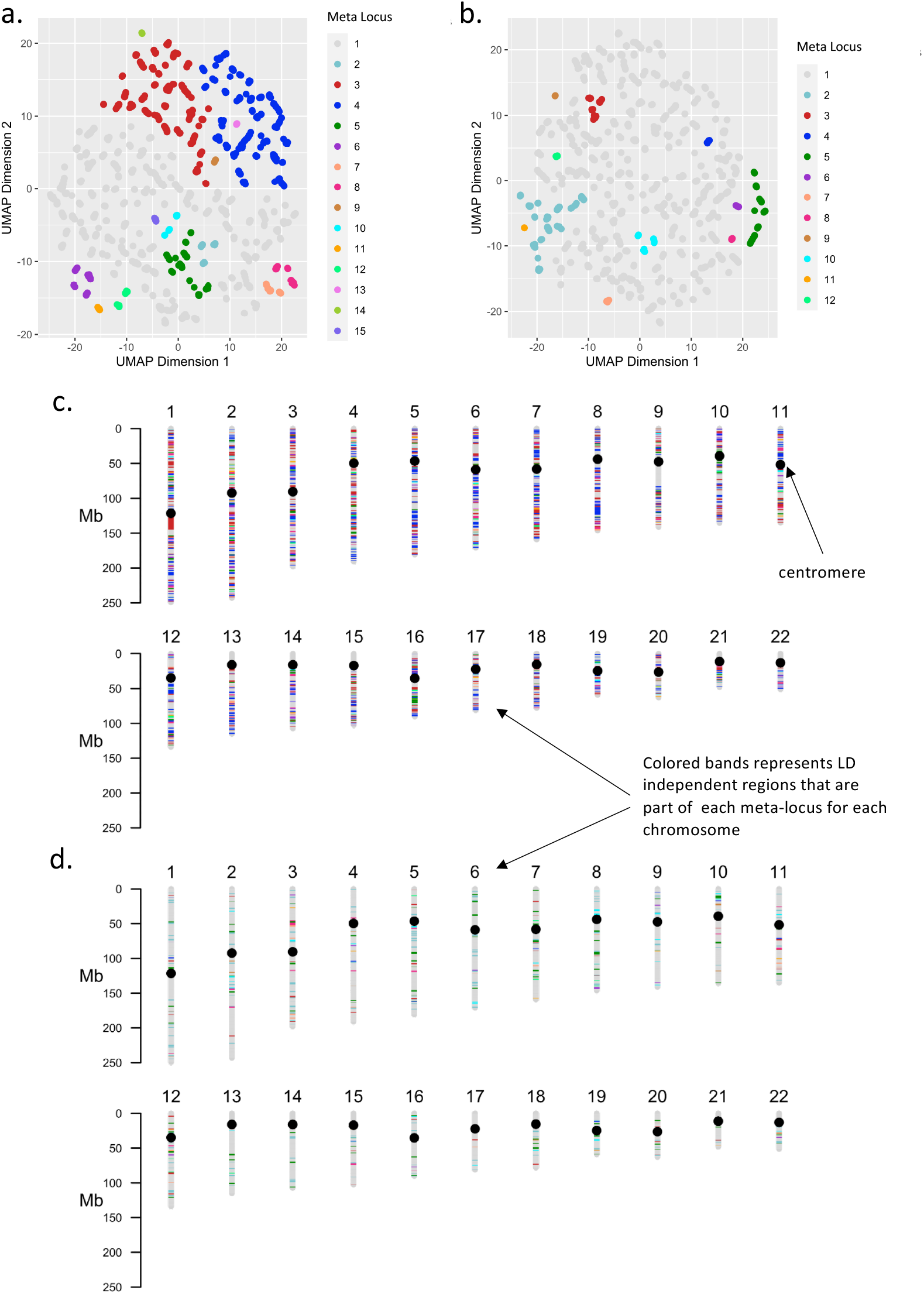
UMAP and Karyogram Plots for Meta-Locus Definitions. *Note*: Panel (a), (b). UMAP plot showing 15 meta-loci and 12 meta-loci identified by Density-based scan procedures across genome-wide LD independent regions for General Cognitive Ability and Non-Cognitive Skills respectively. Panel (c), (d). Karyogram plots for General Cognitive Ability and Non-Cognitive Skills respectively. Color scheme in each of the Karyogram plots matches those reflected in Panels (a) and (b).

The pattern of local genetic correlation for each of the prioritized meta-loci is displayed in Fig-3 (distribution for all meta-loci displayed in Fig-S5 & point estimates of mean local genetic correlation Z-scores displayed in Fig-S6). Schizophrenia and bipolar disorder showed similar local genetic correlation profiles for the prioritized NCS meta-loci but were differentiated on the GCA meta-loci. As expected, Schizophrenia showed strong trends of negative genetic correlation in most GCA meta-loci, with positive local genetic correlations observed in only 2 of the 8 prioritized GCA meta-loci. Notably, attention-deficit/hyperactivity disorder (ADHD) was negatively associated across all GCA and NCS meta-loci, whereas obsessive compulsive disorder (OCD) was positively associated with both cognitive dimensions across all meta-loci. Affective and anxiety related traits showed expected broad negative associations with both cognitive dimensions, but appeared differentiated on specific meta-loci e.g., GCA-3. Autism spectrum disorder (ASD) demonstrated a relatively unique pattern of relationships, with relatively modest effects across most meta-loci, except for a positive genetic correlation with GCA at meta-locus GCA-7, and a negative correlation at NCS-3.

**Figure 3.**
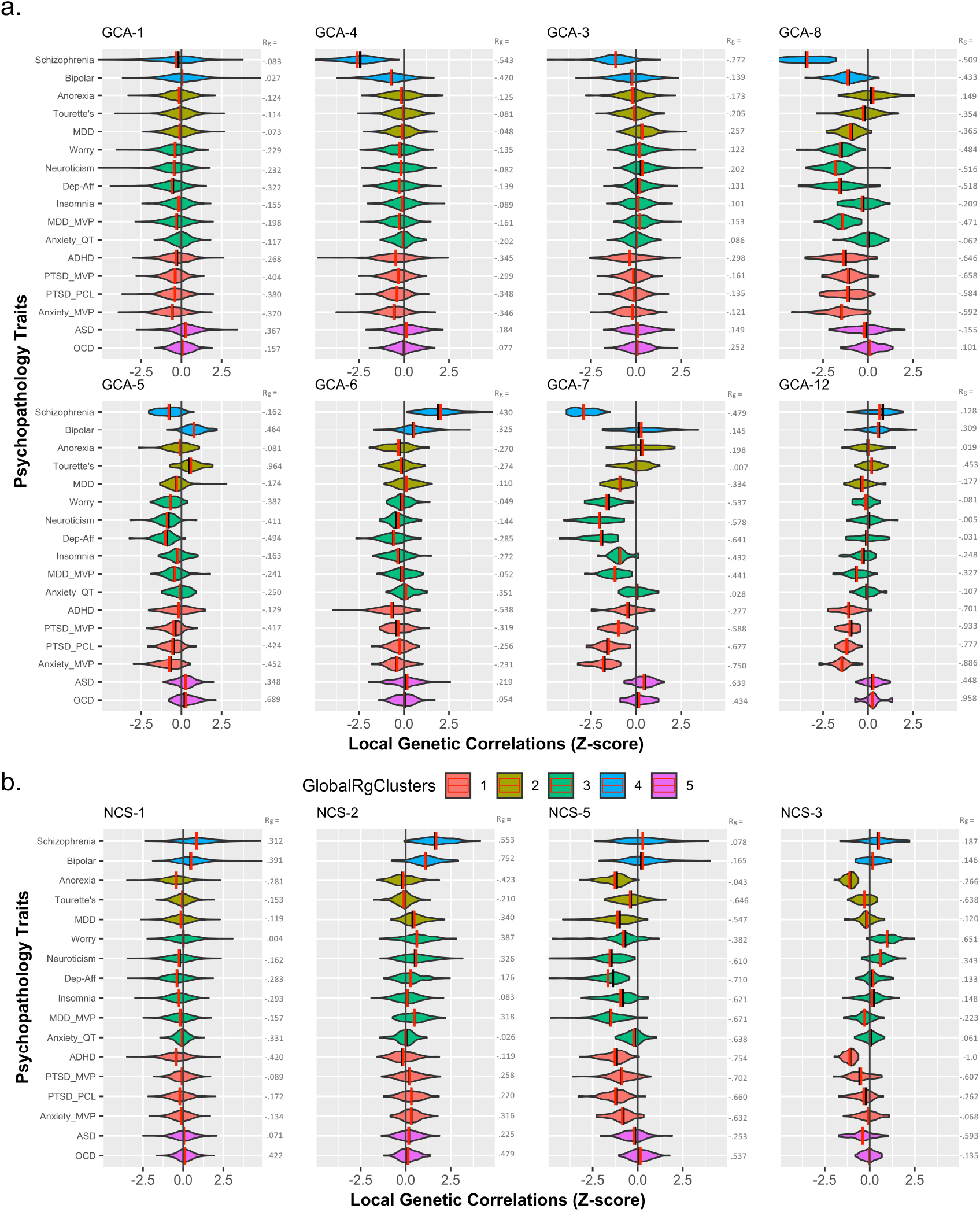
Violin plots for Z-score distributions of Local Genetic Correlations within each prioritized meta-locus. *Note*: GlobalRgClusters: Phenotype clusters derived from partitioned clustering of global genetic correlation. Panel (a). Prioritized meta-loci for General Cognitive Ability. Panel (b). Prioritized meta-loci for Non-Cognitive Skills. Panel (a), (b). Bipolar: Bipolar Disorder, Anorexia: Anorexia Nervosa, Tourette’s: Tourette’s Syndrome, MDD: Major Depressive Disorder (Howard et al., 2019), Dep-Aff: Depressive Affect, MDD_MVP: Major Depressive Disorder (Million Veteran Project), Anxiety_mvp: Anxiety Disorder (Million Veteran Project), PTSD_mvp/pcl: Post-Traumatic Stress Disorder (Million Veteran Project; Total PCL: Total PCL symptom scores)

It was important to determine if the pattern of results was driven by socioeconomic status, given that prior literature^33^ has indicated that educational attainment exhibits shared biology with socioeconomic status. We carried out local genetic correlations of GCA and NCS and the Townsend Social Deprivation Index via ρ-HESS, in a similar manner detailed above. As displayed in Fig-S7/Table-S7, most meta-loci showed negligible association.

#### Stage 3: Functional annotation and gene prioritization for characterizing biology underlying GCA and NCS Meta-Loci

In order to characterize each meta-locus, we first applied a series of gene-based and TWAS methods to the GCA and NCS GWAS summary statistics (gene-based results: Table-S8; TWAS results: Table-S9/S10/S11). As an initial loose filter, genes were ranked based on multiple complementary transcriptome analyses (Table-S12), applying a 50^th^ percentile cut off for gene association p-values; any genes with no evidence of transcriptomic association to the cognitive phenotypes were removed from further consideration. PoPs gene prioritization scores^32^ (Table-S13) for each remaining gene within a given meta-locus were then subjected to gene set analyses, using annotations obtained from the GO Gene Sets (Cellular Component, Molecular Function and Biological Process) included in the Molecular Signature Database v 7.2^34^. Detailed descriptions of the parameters applied to each of the methods are described in the Methodology section. Gene set analyses were carried out via three methods [Gene Set Enrichment Analysis (GSEA^29^), WebGestalt^30^, and GENE2FUNC (part of the Functional Mapping and Annotation of Genetic Association – FUMA – pipeline^31^)];; we conservatively required agreement (Bonferroni-corrected significance) from all three methods in order to designate a gene set as significant for a given meta-locus.

The gene set analysis described above identified a total of 62 gene sets underlying 6 GCA meta-loci (GCA-1,3,4,5,6,8) and 35 gene sets underlying 3 NCS meta-loci (NCS-1,2,5) (Fig-4a, and Table-S14); the remaining meta-loci had no gene sets meeting our strict significance criteria. In Fig-4, several large neurobiological gene sets which were implicated across both GCA and NCS meta-loci are colored red (Fig-4a/b) and those that overlapped multiple GCA meta-loci (but no NCS meta-loci) are colored orange (Fig-4c). The remaining gene sets, which are specific to individual GCA or NCS meta-loci, are colored gray.

**Figure 4.**
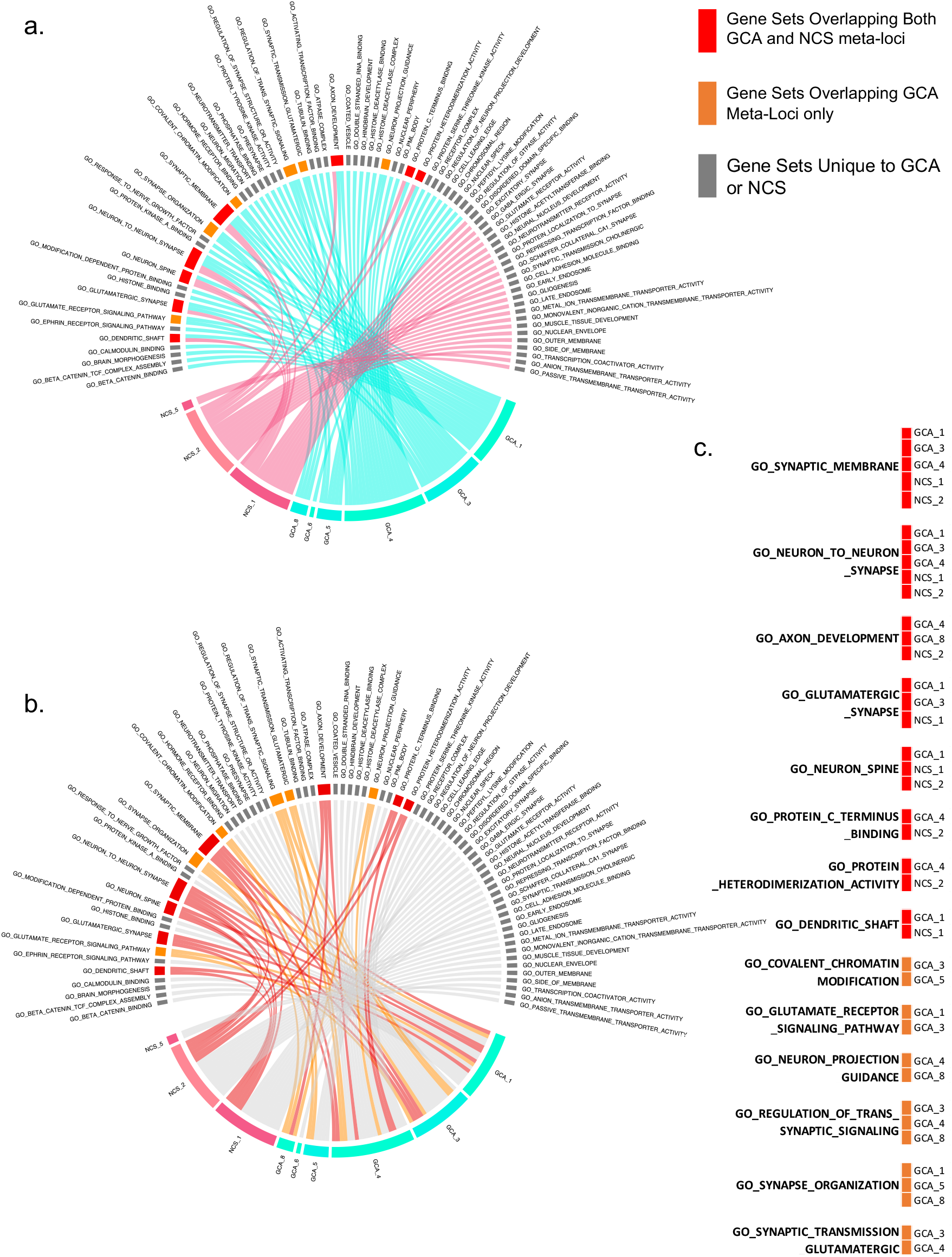
Converging gene sets identified within each General Cognitive Ability and Non-Cognitive Skills Meta-Locus. *Note*: GCA: General Cognitive Ability, NCS: Non-Cognitive Skills. Panel (a), (b), Top half of circular plots are gene sets, bottom half are the meta-loci. Panel (c). Overlapping gene set expanded for easy reference.

Focusing on gene sets specific to individual meta-loci might allow more targeted parsing of the biological mechanisms underlying the relationship between psychopathology and cognition. There were 34 such gene sets across GCA meta-loci, sharing a predominately neurodevelopmental theme as elaborated in Table 1 (e.g., GCA-1: Neurodevelopment/ Neuroplasticity; GCA-4: Neural Connectivity/Differentiation). By contrast, the 24 gene sets specific to individual NCS meta-loci shared a predominant theme relating to synaptic transmission (e.g., NCS-1: Synaptic Activity/Neurotransmission; NCS-2: Synaptic Transmission /Potentiation; NCS-5: GABA-ergic Synaptic Transmission) (Table 2). Pathway definitions for each prioritized meta-locus, as a function of psychopathologic correlations are summarized in Fig-S8.

**Table 1.** General Cognitive Ability – Prioritized Meta-Loci, Gene Sets and Tier 1 Druggable Genes

| GCA-1 | GCA-4 | GCA-3 | GCA-5 | GCA-8* |
| --- | --- | --- | --- | --- |
| $h^2 = 0.104$ (45.8%) | $h^2 = 0.045$ (19.8%) | $h^2 = 0.032$ (13.9%) | $h^2 = 0.009$ (3.9%) | $h^2 = 0.010$ (4.6%) |
| N LD regions = 1125 | N LD regions = 430 | N LD regions = 377 | N LD regions = 83 | N LD regions = 55 |
| Length = 1340.79 (mb) | Length = 514.32 (mb) | Length = 439.17 (mb) | Length = 101.61 (mb) | Length = 73.7 (mb) |
| Neurodevelopment/<br>Neuroplasticity | Neural Connectivity | Synaptic Structure/<br>Neurodegeneration | Neuronal Structure | Axonal Guidance |
| • Histone Binding<br>• Response to Nerve Growth Factor<br>• Beta Catenin Binding<br>• Beta Catenin TCF Complex Assembly<br>• Ephrin Receptor Signaling Pathway<br>• Modification Dependent Protein Binding<br>• Protein Kinase A Binding<br>• Brain Morphogenesis<br>• Calmodulin Binding | • Coated Vesicle<br>• Double Stranded RNA Binding<br>• Activating Transcription Factor Binding<br>• Histone Deacetylase Binding<br>• ATPase Complex<br>• Nuclear Periphery<br>• Protein Serine Threonine Kinase Activity<br>• PML Body<br>• Histone Deacetylase Complex<br>• Regulation of Neuron Projection Development<br>• Receptor Complex<br>• Hindbrain development | • Neuron Migration<br>• Regulation of Synapse Structure or Activity<br>• Presynapse<br>• Neurotransmitter Transport<br>• Tubulin Binding<br>• Hormone Receptor Binding<br>• Protein Tyrosine Kinase Activity<br>• Phosphatase Binding | • Cell Leading Edge<br>• Peptidyl Lysine Modification<br>• Nuclear Speck<br>• Chromosomal Region | • Axon Development<br>• Neuron Projection Guidance<br>• Synapse Organization<br>• Regulation of Trans Synaptic Signaling |
| **Calcium/calmodulin dependent protein kinase:** CAMK1D, CAMK2A, CAMK2G, CAMKK1,<br>**Ephrin Receptor:** EPHA10, EPHA3, EPHA6, EPHA7, EPHB1<sup>Δ</sup>,<br>**Glutamate Receptor:** GRIN1, GRIN2B,<br>**Protein Kinase:** PRKCB, PRKACA,<br>**Others:** ADCY1, APH1A, CACNA1C<sup>Δ</sup>, CDH2, EEF2, GSK3A, HDAC1, KMT2A, LRRK2, MAPT, MKNK2, NTRK3<sup>Δ</sup>, PDPK1, PHKG2, PKN1, PPP3CA, PSEN1, RARA, ROCK1, SIRT1<br>*Total: 33 genes* | **Casein Kinase:** CSNK1D, CSNK2A1,<br>**Ephrin Receptor:** EPHA5, EPHA8, EPHB2<sup>Δ</sup>,<br>**Glutamatergic Receptor:** GRIK2<sup>Δ</sup>, GRIN2A<sup>Δ</sup>,<br>**Histone Deacetylase:** HDAC11, HDAC3,<br>**Mitogen Activated Protein:** MAP2K5, MAP3K10, MAP3K12, MAP3K2, MAP4K4, MAP4K5,<br>**Serine/Threonine Kinase** STK11, STK35,<br>**Other:** ABL1, ACVR1B, AKT2, APP, BRSK2, CDK13, CNR1, DNMT3A, DYRK1B, HIPK1, HSP90AB1, IGF1R, MARK1, NCOR2, NTRK2, PAK1, PKN2, PRKCG, RAF1, ROCK2, RPS6KB1, SHH, SLC6A4, SRPK2, TAOK3, TLK2, TNIK, VEGFA<br>*Total: 45 genes* | **CDC-like Kinase:** CLK2, CLK4,<br>**Ephrin Receptor:** EPHA4, EPHB2<sup>Δ</sup>, EPHB6<br>**Glutamatergic Receptor:** GRIA1, GRIK2<sup>Δ</sup>, GRIN2A<sup>Δ</sup>,<br>**Solute Carrier family:** SLC6A1, SLC9A1,<br>**Others:** AKT1, CAMK2B, CDK5, CSK, ERBB4<sup>Δ</sup>, ESR1, FGFR3, FKBP4, HTT, JAK2, MAPK14, MET<sup>Δ</sup>, NTRK3<sup>Δ</sup>, RXRA, SRC, TP53<br>*Total: 26 genes* | EPOR<br>*Total: 1 gene* | PTK2, GRIK3, PIK3CA, NISCH, EPHA10, NCAM1, VAMP2, DRD2, CACNA1D<br>*Total: 9 genes* |

**Table 2.** Non-Cognitive Skills – Prioritized Meta-Loci, Gene Sets and Tier 1 Druggable Genes

| NCS-1 | NCS-2 | NCS-5 |
| --- | --- | --- |
| $h^2 = 0.244$ (78.3%) | $h^2 = 0.024$ (7.6%) | $h^2 = 0.020$ (6.3%) |
| N LD regions = 1926 | N LD regions = 162 | N LD regions = 89 |
| Length = 2290.00 (mb) | Length = 191.66 (mb) | Length = 116.4 (mb) |
| Synaptic Activity/<br>Neurotransmission | Synaptic Transmission/<br>Action Potentiation | GABA-ergic Synaptic<br>Transmission |
| •Neurotransmitter Receptor Activity<br>•Disordered Domain Specific Binding<br>•Schaffer Collateral CA1 Synapse<br>•Repressing Transcription Factor Binding<br>•Excitatory Synapse<br>•Protein Localization to Synapse<br>•Gaba-ergic Synapse<br>•Neural Nucleus Development<br>•Histone Acetyltransferase Binding<br>•Synaptic Transmission Cholinergic<br>•Glutamate Receptor Activity | •Metal Ion Transmembrane Transporter Activity<br>•Monovalent Inorganic Cation Transmembrane Transporter Activity<br>•Muscle Tissue Development<br>•Early Endosome<br>•Late Endosome<br>•Outer Membrane<br>•Side of Membrane<br>•Cell Adhesion Molecule Binding<br>•Nuclear Envelope<br>•Transcription Coactivator Activity<br>•Gliogenesis | •Anion Transmembrane Transporter Activity<br>•Passive Transmembrane Transporter Activity |
| **Cyclin Dependent Kinase:** CDK5, CDK5R1,<br>**Cholinergic Receptor:** CHRM1, CHRM2, CHRNA1, CHRNA2, CHRNA3, CHRNA4, CHRNA7, CHRNB1, CHRNB2, CHRNB4, CHRNE, CHRNG,<br>**Dopamine Receptor:** DRD1, DRD5,<br>**GABAergic Receptor:** GABRA5, GABRB2<sup>Δ</sup>, GABRB3, GABRG3,<br>**Glutamatergic Receptor:** GRIA1, GRIA2, GRIA4, GRIK2, GRIN1, GRIN2C, GRIN2D, GRM5,<br>**Histone Deacetylase:** HDAC1, HDAC2, HDAC4, HDAC5, HDAC7, HDAC9<sup>Δ</sup>,<br>**Heat Shock Protein:** HSP90AA1, HSP90AB1,<br>**5-hydroxytryptamine receptor:** HTR1B, HTR1F,<br>**Solute Carrier Family:** SLC6A1, SLC6A9,<br>**Others:** ADORA1, BCL2<sup>Δ</sup>, BCR, CNR1, EPHA7, ERBB4<sup>Δ</sup>, FYN, HIF1A, HRAS, MAPT, MDM2, NQO1, PDE4B<sup>Δ</sup>, PPP3CA, PTK2B, SV2A, TP53<br>*Total: 57 genes* | **Apoptosis regulator:** BCL2<sup>Δ</sup>, BCL2L1,<br>**Ephrin receptor:** EPHA2, EPHA4, EPHB1,<br>**Intercellular Adhesion Molecule:** ICAM1, ICAM3,<br>**Other:** ATP1A2, GHR<sup>Δ</sup>, HMGCR, IL11RA, ITGAL, KCNK3, MAPK3, PAK6, PPARG, PTPRC, RAF1, SIGMAR1<br>*Total: 19 genes* | **GABAergic Receptor:** GABRA1, GABRA2, GABRA4, GABRA6, GABRB1, GABRB2<sup>Δ</sup>, GABRG1, GABRG2, GRIN2B,<br>**Other:** KCNB2, P2RX4, SLC12A5, SNAP25<br>*Total: 13 genes* |

There were 906 GCA and 493 NCS “driver” genes identified from the GSEA analysis. The reduced list of genes was further examined if they were potentially actionable in terms of having encoding proteins of putative drug targets, using chemoinformatic annotations provided by Finan et al. ^35^ (Table-S15). Tier 1 druggable genes (i.e., genes with current evidence of having existing compounds that are FDA approved and being utilized for various indications), summarized alongside other meta-locus information, are listed in Table 1 and Table 2. Several gene families with known druggable components were identified, including several glutamatergic and ephrin receptor genes identified within GCA meta-loci. Most notably, the NCS-5 meta-locus implicates multiple GABA receptor genes across disparate chromosomal regions, suggesting the possibility that GABA-ergic treatment approaches may enhance NCS while simultaneously ameliorating the correlated affective and anxiety symptoms that load on this meta-locus.

The dichotomy between neurodevelopment and synaptic pathways underlying GCA and NCS, respectively, suggested an additional hypothesis to be tested. If GCA was primarily driven by neurodevelopmental genes, we would expect to see gene expression profiles that are active prenatally or early in the lifespan, whereas for NCS, genes responsible for synaptic transmission are likely to be expressed later in postnatal life. Leveraging the BrainSpan dataset^35^ which characterized brain expression profiles at various developmental stages, we tested if GCA and NCS driver genes might demonstrate differential expression across the lifespan. By fitting a linear mixed model, with individual as random effect, cognitive phenotypes and time as fixed effects and sex as a covariate, we found developmental differences between GCA and NCS driver genes (for further details, see Online Methodology and Supplementary Information). Setting the null model simply with time as predictor, we observed a significant improvement in model fit when an indicator of the cognitive phenotype (GCA vs. NCS) for the driver gene was included in the model (χ^2^ = 1059.6, df = 2, p = 8.14e-231). A main effect for differences in GCA and NCS genes was found (t = 10.64, df = 7, p = 1.42e-5), and an interaction between GCA and NCS genes with time (t = 28.28, df = 7, p = 1.78e-8) was also observed (Fig-5, Fig-S9). As hypothesized, genes harbored within NCS-meta loci were expressed predominantly in early adulthood and adulthood, whereas GCA-meta loci genes were expressed pre-natally.

**Figure 5.**
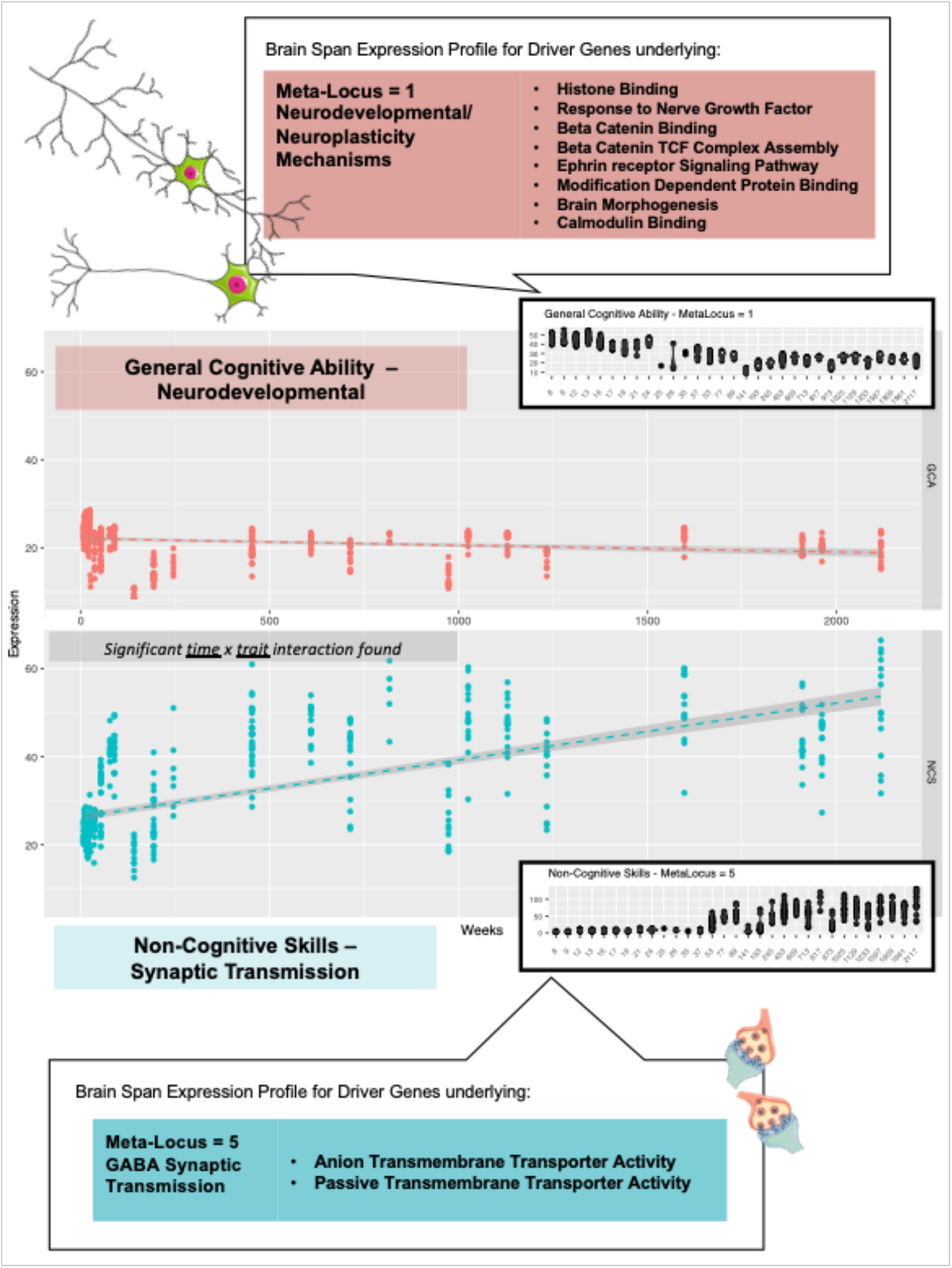
BrainSpan gene expression profiles for General Cognitive Ability and Non-Cognitive Skills. *Note*: GCA: General Cognitive Ability, NCS: Non-Cognitive Skills. Panel (a) scatterplot for GCA (b) scatterplot for NCS. Y-axis: Normalized gene expression x-axis: Time in weeks.

## Discussion

Numerous recent studies have demonstrated shared genetic underpinnings amongst multiple forms of psychopathology, as well as between psychiatric and cognitive phenotypes^10,11,23,37^. The present study was designed to parse this genetic overlap into separable biological pathways with specific psychiatric and cognitive subcomponents. The present study extends our previous work, in which we leveraged cognitive pleiotropy to differentiate early neurodevelopmental mechanisms from synaptic function in the etiopathogenesis of schizophrenia^15^. Here, we demonstrated that prenatal neurodevelopmental mechanisms shape the relationship between general cognitive ability and multiple forms of psychopathology, while synaptic pathways expressed later in life underlie the paradoxical association between enhanced non-cognitive skills and greater risk for psychotic disorders. Prior studies of pleiotropy^10,11,14^ have implicated neurodevelopmental and synaptic pathways as shared components underpinning multiple psychiatric disorders, but without the ability to differentiate these mechanisms (Table-S16). By contrast, gene set analysis of individual trait GWAS in psychiatry (Table-S16) have either failed to identify one^38,39^ or both^40^ of these pathways, or have implicated these pathways without directionality or subsetting^41,42^

Within the broad neurodevelopmental and synaptic themes, we were able to further specify individual gene sets with distinct patterns of association to psychiatric and cognitive phenotypes. These discoveries were made possible by our novel construction of “meta-loci”, defined by the concatenation of LD-independent genomic regions with shared patterns of local genetic correlations across phenotypes. Within meta-loci, we further identified genes that may serve as actionable targets, or at least actionable entry points into relevant biological pathways, for psychiatric and nootropic drug repurposing (Table 1 and Table 2); several notable examples are described below.

The GCA-8 meta-locus spans 55 LD-independent regions, covering the genomic length of 73Mb, and is characterized by “Axon Development” and related gene sets. We found that this meta-locus shows the strongest pleiotropy with schizophrenia, but also implicates bipolar disorder and a series of affective phenotypes. Notable driver genes in this meta-locus included *DRD2, NCAM1, DCC, CACNA1D*, and *SLC4A10*, among many known genes that implicate psychopathology. In their most recent report, the PGC Cross Disorder group reported that the *DCC* region was most pleiotropic in psychopathology. The *DCC* gene was a known master regulator of early neurodevelopmental biology via interactions with netrin-1 and draxin, and has been implicated in developmental processes of white matter tracts in the brain^43,44^. In the report, the authors suggested that the *DCC* is likely to affect childhood developmental disorders such as ADHD and ASD, which they have demonstrated to cluster together. Importantly, our evidence showed that similar processes are not limited to neurodevelopmental disorders but are implicated in other psychopathological phenotypes and are linked through a shared effect on general cognitive ability.

By contrast, the NCS-5 locus showed the strongest trend for adulthood spatial-temporal gene expression and the driver genes were primarily inhibitory GABA genes, several of which were previously implicated in major depressive disorder^45^ and post-traumatic stress disorder^46^. Similarly, driver genes at the NCS-2 meta-locus, characterized by “Synaptic Transmission” and related gene sets, showed predominant postnatal/adulthood expression. This meta-locus shows strong pleiotropy with psychotic disorders (schizophrenia and bipolar disorder); to lesser extent positive local genetic correlations were also observed in affective disorders and anxiety. *AKAP6, CADM3*, and *PDE4B* were notable genes implicated in both schizophrenia and bipolar in earlier reports^47–49^. At the same time, novel associations were observed to *PPARG* and *HMGCR*, both of which are critical to metabolism and energy utilization, possibly suggesting an unexplored mechanism of neuronal function. Given the counterintuitive pleiotropy at this locus, with greater non-cognitive skills associated with increased risk for psychopathology, such mechanistic leads may prove fruitful for further experimentation.

The meta-locus approach also extends and refines prior work on the structure of psychopathology. Recently, both the PGC Cross Disorder Group^10^ as well as Grotzinger and colleagues^36^ utilized global genetic correlations to show converging evidence for several latent factors underlying current nosological constructs in psychiatry: (i) Psychosis factor (schizophrenia and bipolar disorder); (ii) Neurodevelopmental factor (alcohol use, ADHD, ASD, PTSD); (iii) Compulsive factor (anorexia nervosa, OCD, and Tourette’s Syndrome); and (iv) Internalizing factor (MDD, anxiety disorders). In this report, schizophrenia and bipolar disorder demonstrated similar patterns of pleiotropy globally, but distinctions were observed at several meta-loci (e.g., GCA-5 and GCA-7), where schizophrenia more closely resembled major depression and other affective phenotypes. Overall, schizophrenia harbors LD-independent segments strongly associated with either GCA or NCS beyond that of other psychopathological phenotypes. Though the genetic correlation between schizophrenia and cognitive dimensions appears modest (r_g_ ≈ -0.2) at the global level, this may reflect mutually negating effects at the regional level across the genome. Broad pleiotropic profiles for GCA/NCS within ADHD, PTSD, MDD, Anxiety disorder and Tourette’s Syndrome support cross factor loadings observed by Grotzinger and colleagues^36^ within “Internalizing” and “Neurodevelopmental factors”. However, in the present study, ADHD and ASD exhibited quite different local genetic correlation profiles across most meta-loci – ADHD being negatively correlated across all meta-loci for GCA and NCS, while ASD demonstrated null or positive correlations with both GCA and NCS. We also demonstrated that the strong positive genetic correlations reported previously between the “Compulsive factor” and Education Attainment was likely accounted for by higher-than-expected genetic correlations between OCD and Non-Cognitive Skills.

Although the MHC region was excluded from most of the downstream work due to its complicated LD patterns, it is worth mentioning that it showed stronger GCA-psychopathology local genetic correlations than other genome regions. Also noteworthy is that Anorexia Nervosa and Tourette’s syndrome showed opposite local genetic correlations within the MHC region relative to all other GCA/NCS - psychopathology trait pairs. Evidence points to the MHC region as potentially a vital aspect of etiopathogenesis in psychopathology. The MHC region harbors the strongest genome-wide signal to date for psychotic and affective disorders. It also harbors a known synaptic pruning mechanism as part of the *C4* complex. The strong association with cognitive ability makes MHC a candidate region for further extensive investigation.

### Conclusions

To conclude, we have leveraged the phenomenon of pleiotropy between cognitive dimensions and psychopathology to dissect underlying biological mechanisms. We demonstrated that beyond broad genome-wide relationships, specific regions of the genome are likely to harbor biological processes that act differentially across psychopathological traits. The phenomenon is unveiled through the combined local genetic correlation analysis and “meta-locus” strategy adopted within this report. Follow up efforts such as increasing the power of univariate traits and the accuracy of reference panels may increase the resolution of such approaches. Also, it seems necessary to devise a way in which latent factors such as Non-Cognitive Skills derived purely from genetic correlations and GWAS summary statistics could be operationalized and measured tangibly in clinical populations. Finally, it should be noted that the present study utilizes well-powered, currently available GWAS that are based on common genetic variation and community sampling; in the future, well-powered family based GWAS may ultimately provide more accurate estimates of genetic effects, including rare variation^50^, independent of “genetic nurture”^51^. Nonetheless, the results reported currently underscores the need to rethink how rapidly available multivariate data within cognitive and psychiatric genomics could be leveraged to uncover more in-depth biological insights beyond simply pointing towards neuronal mechanisms.

## Online Methodology

### Genome-Wide Summary Statistics Quality Control and Harmonization

The first stage of the analysis involved the curation of the GWAS summary statistics of cognitive features and psychopathological traits. We examined the recent literature and selected GWAS of 19 cognition related traits as well as 17 psychopathological traits previously known to be with associated with cognitive function (See Figure 1, Supplementary Table 1). All summary statistics were harmonized based on alleles and frequencies in the 1000 Genomes phase 3 v5 reference^52^. Variants with allele frequencies less than 0.5% and imputation quality less than 0.3 were excluded from the analysis. We also performed allele alignment to resolve strand and allele flips. All summary statistics were aligned based on the A1 allele (reference allele) of the 1000G reference panel^52^. Variants that could not be aligned to the 1000 genomes were excluded. In addition, we excluded ambiguous variants with an allele frequency difference of greater than 0.35 from the reference panel and non-ambiguous variants with an allele frequency difference of 0.15 difference from the reference. Further details are reported in Supplementary Table 1.

### Global Genetic Correlations and Clustering Analysis

We carried out global genetic correlation analysis between the set of 19 cognitive traits and 17 psychiatric traits via LD score regression^21,22^ implemented in Genomic SEM^11^. We followed standard procedures within Genomic SEM as a wrapper for extracting the global genetic covariance matrix (https://github.com/GenomicSEM/GenomicSEM/wiki/). GenomicSEM performs initial data alignment to the HAPMAP3 SNPs and conducts pairwise LD score regression with each pair of input phenotypes for the estimation of global genetic correlations. The global genetic correlations between all cognitive and psychopathological traits were organized into a 19 × 17 matrix and subsequently used as the input for Principal Components Analysis (PCA) and various clustering analyses reported subsequently. The first two principal components were extracted from the 19 × 17 matrix representing the relationship between each psychopathology trait and the top two cognitive dimensions (Table-S2). The loading of each cognitive trait on the principal component was estimated by performing bivariate Pearson correlation between the genetic correlation profile of each cognitive trait and each principal component (columns U and V in Table-S2). The dissimilarity matrix estimated based on Euclidean distance from the global genetic correlation matrix was used to generate the partitioned k-medoid clusters. The clustering procedures were carried out using fviz_cluster() in R as part of the “factoextra” package (https://www.rdocumentation.org/packages/factoextra/versions/1.0.3)

After closely reviewing the global genetic correlation profiles for cognitive features and psychopathological traits described above, GCA and NCS emerged as two candidate traits that were fairly well differentiated across clusters of psychopathological traits. Partitioning cluster analysis (K-medoids and K-means) and hierarchical clustering were employed to examine the latent genome-wide genetic architecture of psychopathological traits in relation to cognitive features. The strategy for the clustering analysis was as follows - we started with a 2-cluster solution, and gradually increased the number of clusters until any of the method reached a singleton cluster. Across methodologies, a 7-cluster solution resulted in a singleton cluster, hence we set the maximum number of clusters to 6 and the minimum number of clusters 2. To select the optimal cluster solution that best represents the data, the procedures detailed in Akhanli and Hennig^53^ were used, which compared model fitting statistics across clustering solutions, using R packges cqcluster.stats() and clusterboot() with default parameters. The best fitting cluster solution was then selected for visualization.

### Data Reduction of Cognitive Features

We applied the GWAS-by-subtraction^16^ procedures developed recently to the latest and largest GCA GWAS^20^ to derive a updated NCS GWAS. We used the Education Attainment GWAS without 23andMe data (N = 495,552), which was slightly smaller compared to the Education Attainment GWAS used in Demange et al.^16^ (N = 565,884). However, our GCA GWAS (N = 373,617) was much more powerful than the one reported previously^16^ (N = 257,841). Nonetheless, LD score regression checks indicated that both GCA and NCS GWAS were highly genetically concordant to previous results (Rg = 1).

### Identify pleiotropy between Cognition and Non-Cognitive Skills and Psychopathological Traits via Local Genetic Correlations

Local genetic correlation analysis was conducted via ρ-HESS^18^. On the advice of the HESS developers, we used a conservative approached that specified full sample overlap between all trait pairs to handle potential unknown sample overlaps. Independent LD segments were estimated using the 1000 genome phase 3 reference panel via LDetect^54,55^. There were a total of 2353 independent LD segments where local genetic correlations for GCA and NCS were carried out vis-a-vis psychopathological traits.

### Identification of Meta-loci via Density-Based Clustering

In order to uncover homogeneous local genetic correlation patterns (between GCA and psychopathological traits and between NCS and psychopathological traits) across the genome, we carried out density-based clustering^56^ in two steps. First, we performed a data reduction step via UMAP^57^ for all pairs of the 19 cognitive features and 17 psychopathological traits across 2353 independent LD segments. Thereafter, density-based scanning was carried out on the first two UMAP components to identify sets of LD independent regions that had homogeneous local genetic correlation profiles between GCA*/*NCS and psychopathology; these sets of LD independent regions were termed as meta-loci. The density-based scan was carried out by first setting the minimum number of LD-independent regions per set as 5 and setting the neighborhood radius to a sufficiently large value such that only one cluster existed. We then reduced the neighborhood radius in increments of 0.1, until the density-based scan yielded a cluster solution *d* with a singleton LD region outlier that could not be clustered. The cluster solution *d* – 1 was used (see Supplementary Information for further details).

### Functional Prioritization of Putative Genes within Meta-loci

To obtain biological insights from the meta-loci we identified, we first prioritized genes within the broad genomic regions based on their possible associations with cognitive phenotypes. We applied a series of gene-based statistical approaches to the GCA and NCS summary statistics derived from the Genomic SEM procedure; these included MAGMA^28^, PoPs^32^, S-Predixcan^58^, SMR/HEIDI^59,60^, and FOCUS^61^. As our goal was to identify relevant genes that may include those below the level of strict genome-wide significance, our initial filter was loose: any gene that was below the 50^th^ percentile of statistical significance for *all* of the above methods was eliminated from further consideration.

For the MAGMA gene-based analysis (v1.08^28^), gene boundaries definitions and the 1000 genomes phase 3 reference panel were provided with the MAGMA package (https://ctg.cncr.nl/software/magma). PoPs allows additional weighting of MAGMA gene-based results, by leveraging the extraction of gene features from comparable genes across all other chromosomes; we applied default parameters as described in Weeks et al.^32^. S-PrediXcan estimates gene-based effects by re-weighting SNP effects with eQTL effects derived from GTEx v8 brain tissue expression data (with LD information derived from the 1000 Genomes phase 3 reference panel). SMR/HEIDI leverages on Mendelian Randomization approaches for gene prioritization; each SNP was treated as an instrument variable, with GCA or NCS as an outcome. Brain expression eQTL data from Brain-eMeta^25^, and PsychENCODE^26,27^ reference datasets are the intermediate variables. As part of the SMR analysis, HEIDI attempts to deconvolve the association to establish if the relationship between SNP, gene expression, and cognitive phenotype are indeed due to modelled causality, or based simply on linkage; SNPs with significant evidence of linkage effects were eliminated from further consideration. FOCUS (Fine-Mapping of Causal Variants)^61^ uses GCA/NCS GWAS summary statistics, expression prediction weights (as estimated from GTEx v8 brain expression reference panels), and LD among all SNPs to estimate the probability for any given set of genes to explain transcriptome association. Two sets of gene ranks emanated from the procedures carried out (i) MAGMA/PoPs (ii) S-PrediXcan/SMR/HEIDI. To prioritize genes, the 50th percentile rank cutoff for (i) & (ii) was stipulated. The final set of filtered genes for gene set analysis was the union of (i) & (ii) and the union of all credible genes identified by FOCUS.

### Elucidating Biological Mechanisms underlying Meta-Loci via Gene Set Analysis

Three separate gene set analysis approaches were then applied to the remaining, filtered gene list for each meta-locus (i) Broad Institute Gene Set Enrichment Analysis (GSEA^29,62^) (ii) WebGestalt^30^ (iii) FUMA GENE2FUNC^31^. GO ontologies within the Molecular Signature Database 7.2 (Biological Processes, Molecular Function and Cellular Component^34,63^) were used as gene-set analysis annotations. Default parameters were used for GSEA and WebGestalt. Both GSEA and WebGestalt require gene scores to be included in the gene set analysis; we utilized the gene prioritization scores derived from PoPs, with an inverse rank transformation applied to the POPs data such that the lowest value of the gene prioritization scores was 0.

Of the three gene set analysis methods, GSEA was arguably the most robust. GSEA first walks down the ranked list of genes, increasing a running-sum statistic when a gene is in the gene set and decreasing it when it is not. The enrichment score is the maximum deviation from zero encountered during that walk. Details of the GSEA methodology has been reported elsewhere^29^. This is followed by WebGestalt^64^ which runs a modified version of GSEA via a web portal. Both GSEA and WebGestalt allow permutation testing for the selection of gene sets which in this case was set to n = 1000 permutations. GSEA analyses were based on the “preranked” procedure, where enrichment scores were normalized. Default filter parameters for minimum (> 15) and maximum (< 500) gene set size were used. MSigDb version 7.2 gene set definitions^34^ for Gene Ontology were used as indicated above. WebGestalt uses a more specifically curated “noRedundant” set of Gene Ontologies based on the 2017 data freeze of the MSigDb. In addition, the method relaxes gene set sizes to permit minimum (> 5) and maximum (< 2000). We set significance to the top 50 gene sets to be extracted for WebGestalt. FUMA GENE2FUNC uses a hypergeometric approach to gene selection which relies only on the overrepresentation of gene symbols for the identification of gene sets. For FUMA GENE2FUNC we set a minimum of 3 genes per gene set and FDR < 0.05 for a gene set to be significant. To select candidate gene sets for each meta-locus, we required a strict “consensus” approach. We deem a gene set as “candidate” only if all three methods identify the same gene set.

In addition to an enrichment score for the gene set, GSEA also generates a set of genes that deviates most from the null during the estimation of the gene set enrichment score. These genes are defined as “driver” genes, because they contribute most to the enrichment of a given gene set. For the purpose of the current report, the driver genes were thus most interesting, as they are likely to be the core genes within a biological pathway.

### *Follow-up Annotations and Analysis of Driver Genes identified within GCA and* NCS *Meta-Loci*

To further understand how driver genes from each gene set might be expressed over time, we performed analyses via the BrainSpan database (based on RNA-Seq Gencode v10 normalized expression values summarized to genes in brain tissues from donors of different ages, ranging from pre-natal to adulthood)^35^. For our analysis, the time of tissue sampling in the BrainSpan database was converted to weeks. All post-natal stages were converted to weeks using *Weeks = Years*52Weeks + 37 Gestational Weeks*. We used linear mixed effects models to investigate if there were significant differences in the gene expression profiles over time between GCA vs. NCS driver genes. The null model included Individual subjects as random effects, and *Time (Weeks)* as a fixed effect predictor; sex was modeled as a covariate. The alternative model additionally included the *Time* (Weeks) *x Trait* (GCA vs. NCS) interaction terms as fixed effect predictors.

Lastly, we examined the potential druggability of the driver genes identified by earlier functional prioritization and gene sets. We extracted data for gene druggability (2047 genes) from Finan et al. ^35^ which included Tier 1, 2, 3A and 3B druggability as well as small molecule druggability, druggability by enzymes or mono-clonal antibodies and drug absorption characteristics. The druggability information was merged with the information available for driver genes identified in the current study. We then further filtered the final list of genes by Tier 1 druggability for display and reporting.

## Data Availability

GWAS summary statistics for GCA and NCS will be made publicly available upon acceptance of the manuscript. Publicly available GWAS summary statistics for psychopathology traits can be obtained from the respective research groups/consortia

## Acknowledgments

Research reported in this publication was supported by the National Institute of Mental Health of the National Institutes of Health under award number R01 MH117646 (TL, Principal Investigator). The content is solely the responsibility of the authors and does not necessarily represent the official views of the National Institutes of Health. WDH and CX are supported by a Career Development Award from the Medical Research Council (MRC) [MR/T03052/1] for the project titled: “From genetic sequence to phenotypic consequence: genetic and environmental links between cognitive ability, socioeconomic position and health.”

## Author Contributions

ML ran the analyses and drafted the manuscript. ML and TL conceived the idea and designed the study. TL supervised the drafting and analyses reported in the manuscript. TG, C-YC, WDH, CX, and RT provided guidance on data analytic approaches. HH, AKM, and HR provided guidance on interpretation of the results. C-YC, ASH, DFL, JG, and MBS provided summary statistics for various GWAS entered into the study. All authors read and provided scientific feedback and participated in finalizing the draft of the manuscript.

## Code Availability

Wrapper for performing local genetic correlations reported in the current report will be made publicly available upon acceptance of the manuscript.

## Statement of ethics

GWAS summary statistics utilized in the current study are publicly available except for the UK Biobank GWAS summary statistics for cognitive phenotypes and the Townsend Index for Social Deprivation which was approved for use under UK Biobank Approved Application ID 26041 (“Large-Scale Sequencing in the UK Biobank to Facilitate Gene Discovery, Genome Sciences, and Precision Medicine”).

