## Supplementary figures and images for "Dissecting Biological Pathways of Psychopathology using Cognitive Genomics"

### FigS1

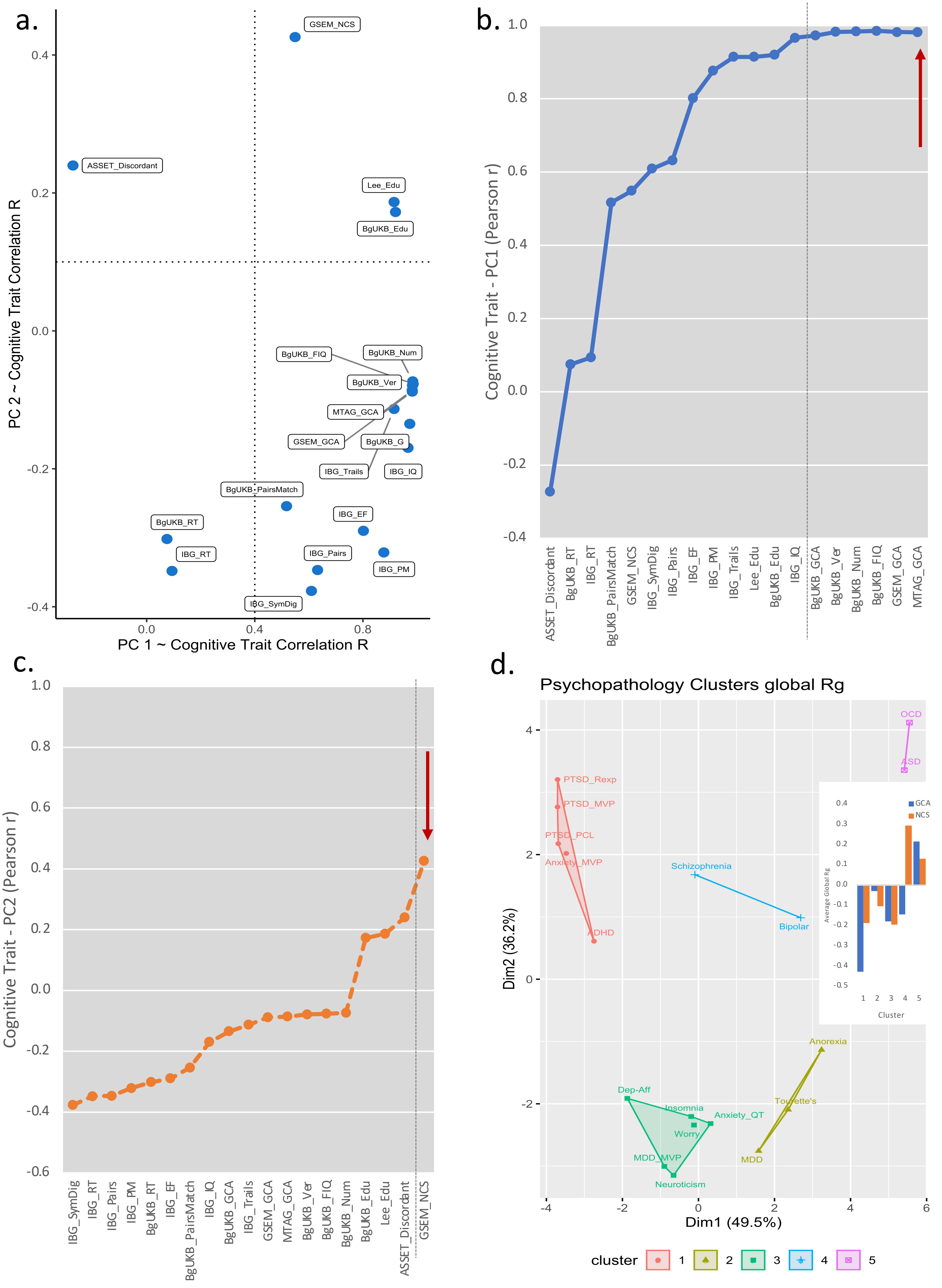

### FigS2

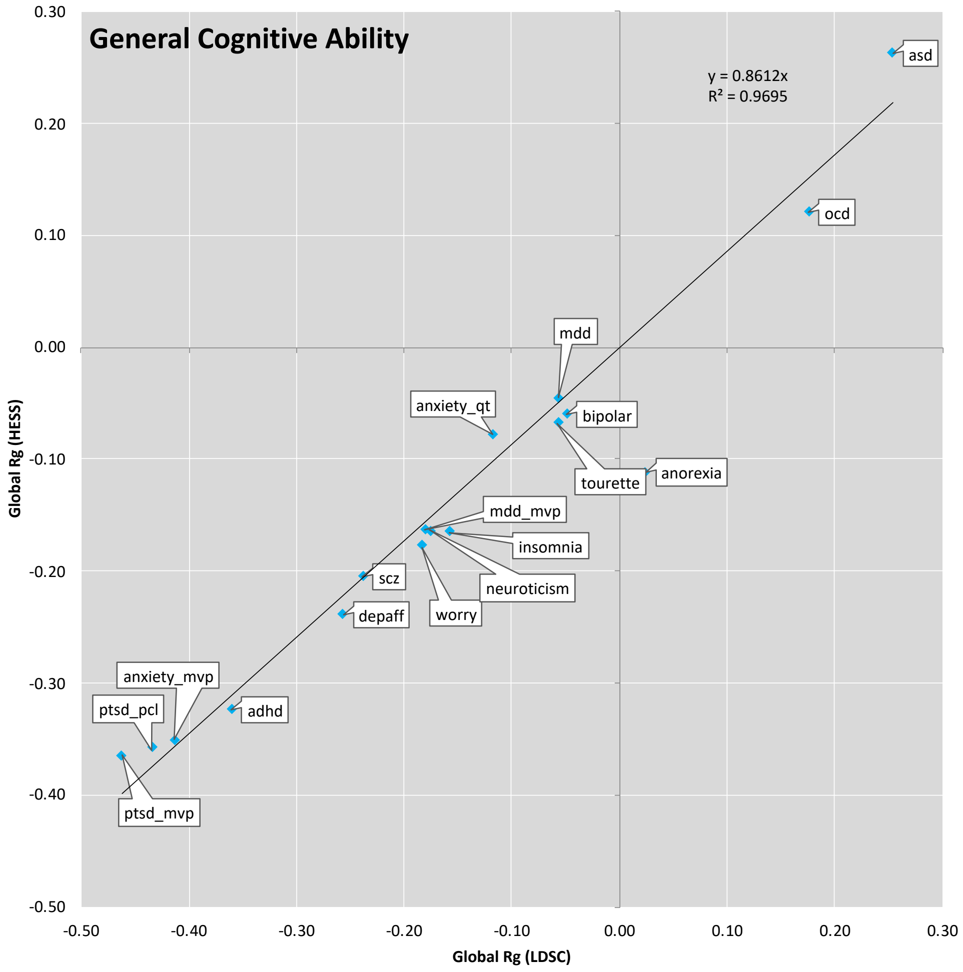

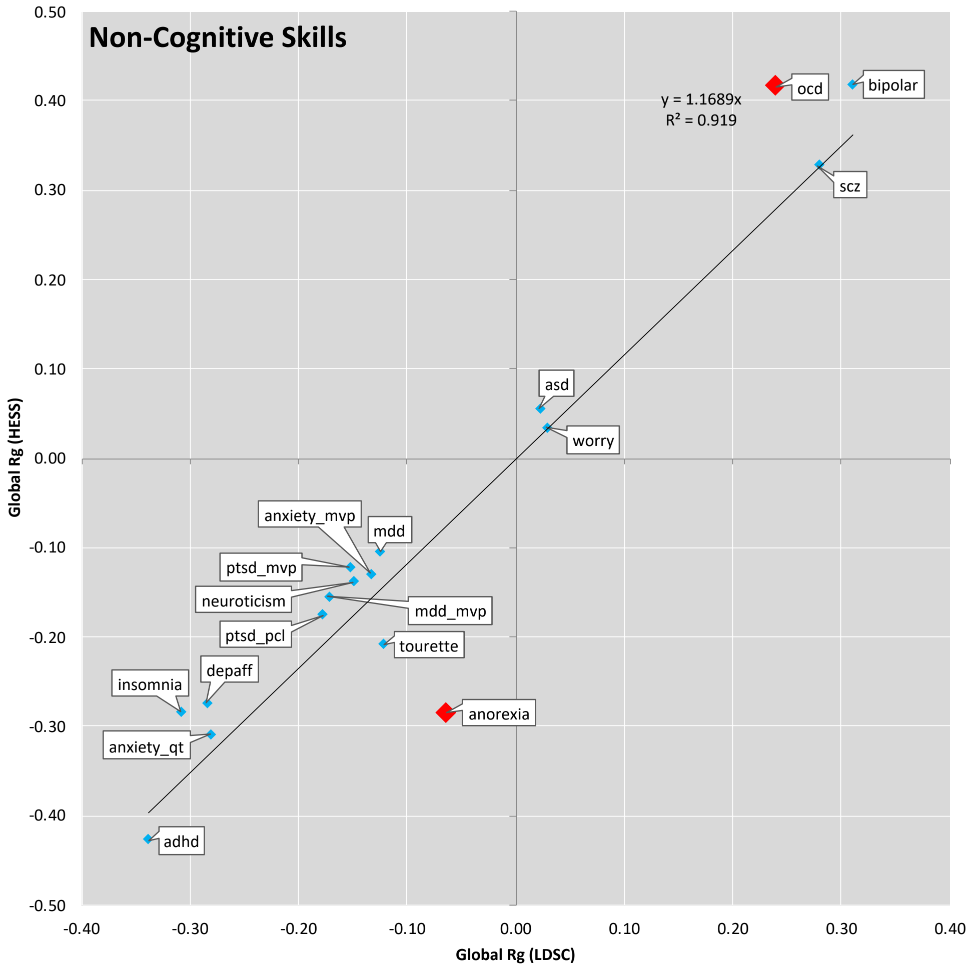

### FigS3

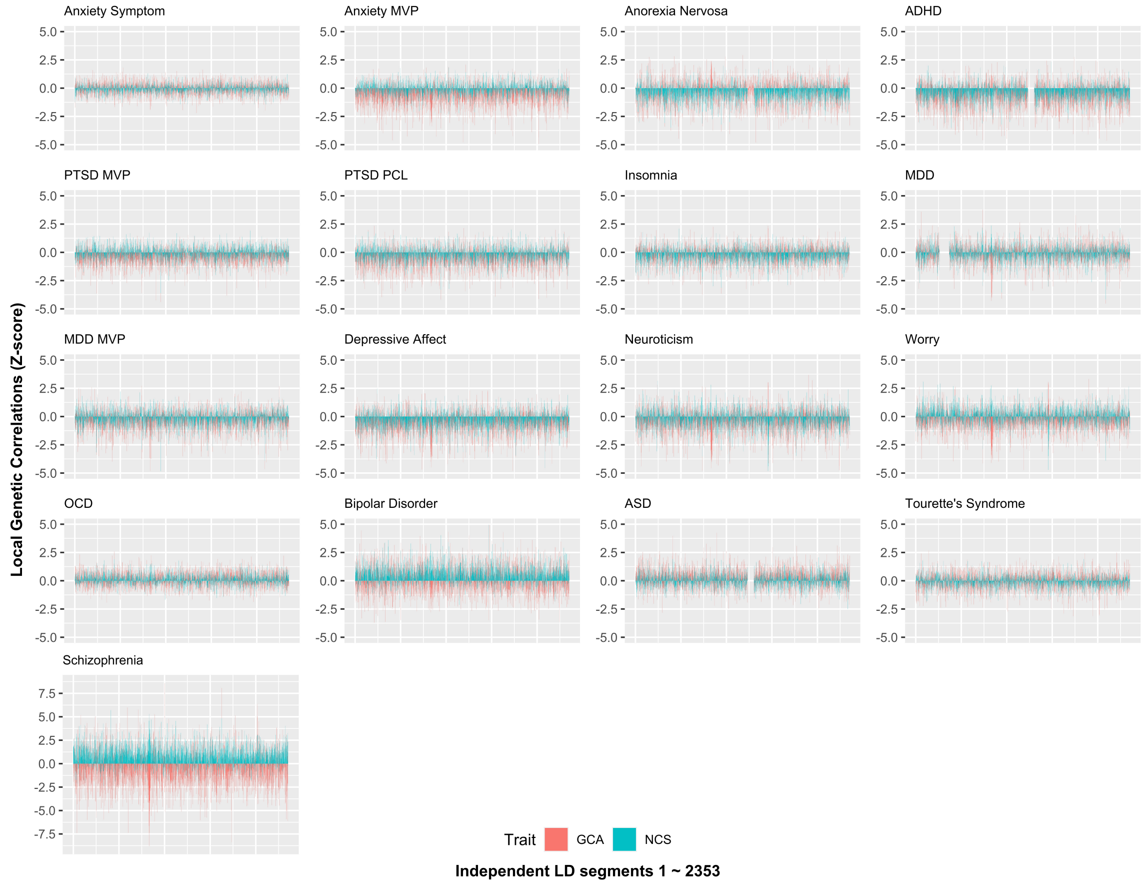

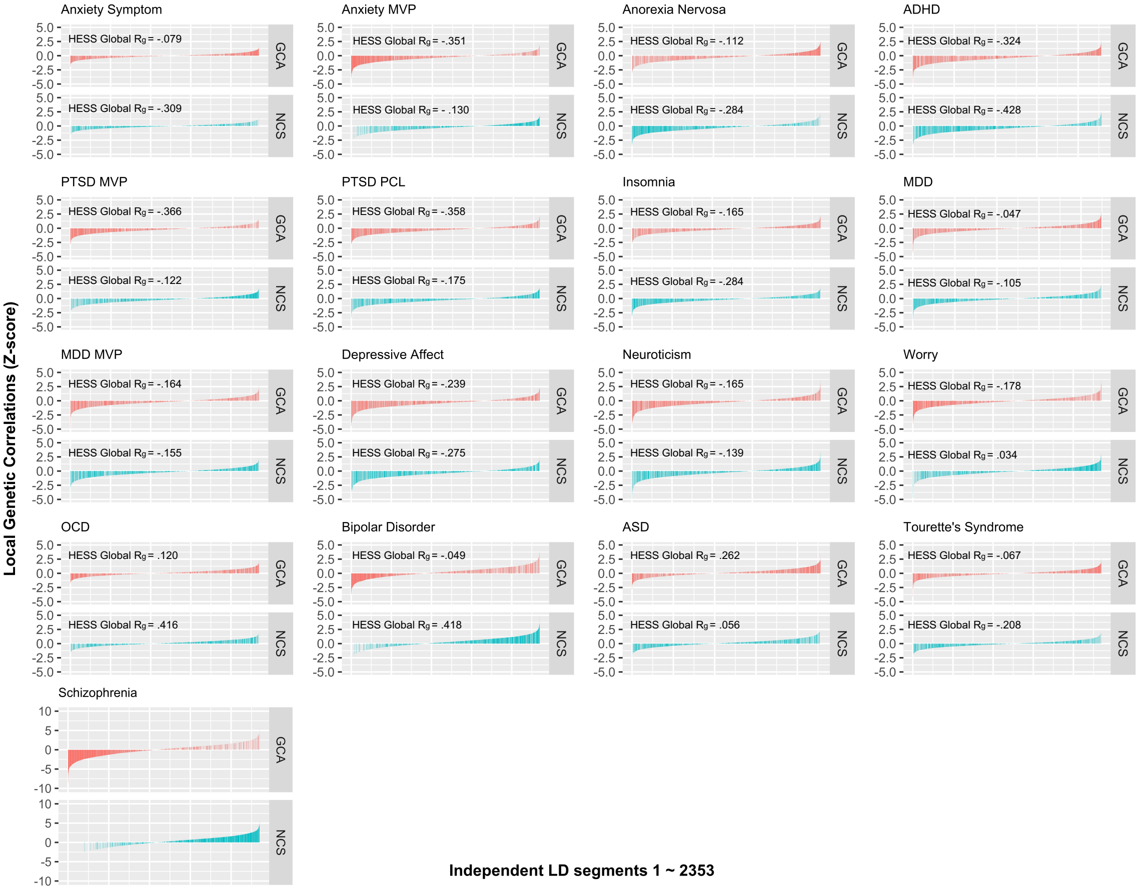

### FigS4

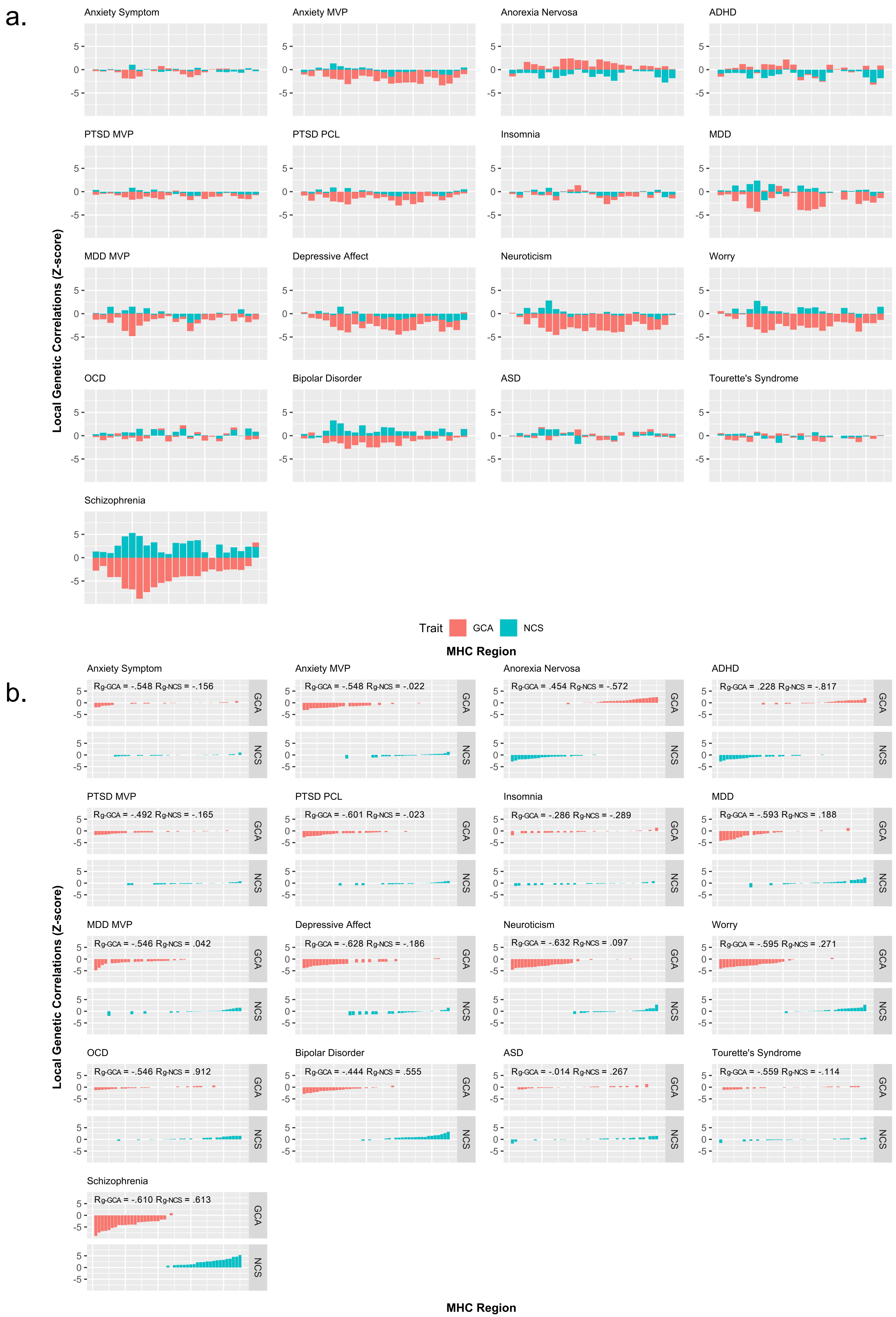

### FigS5

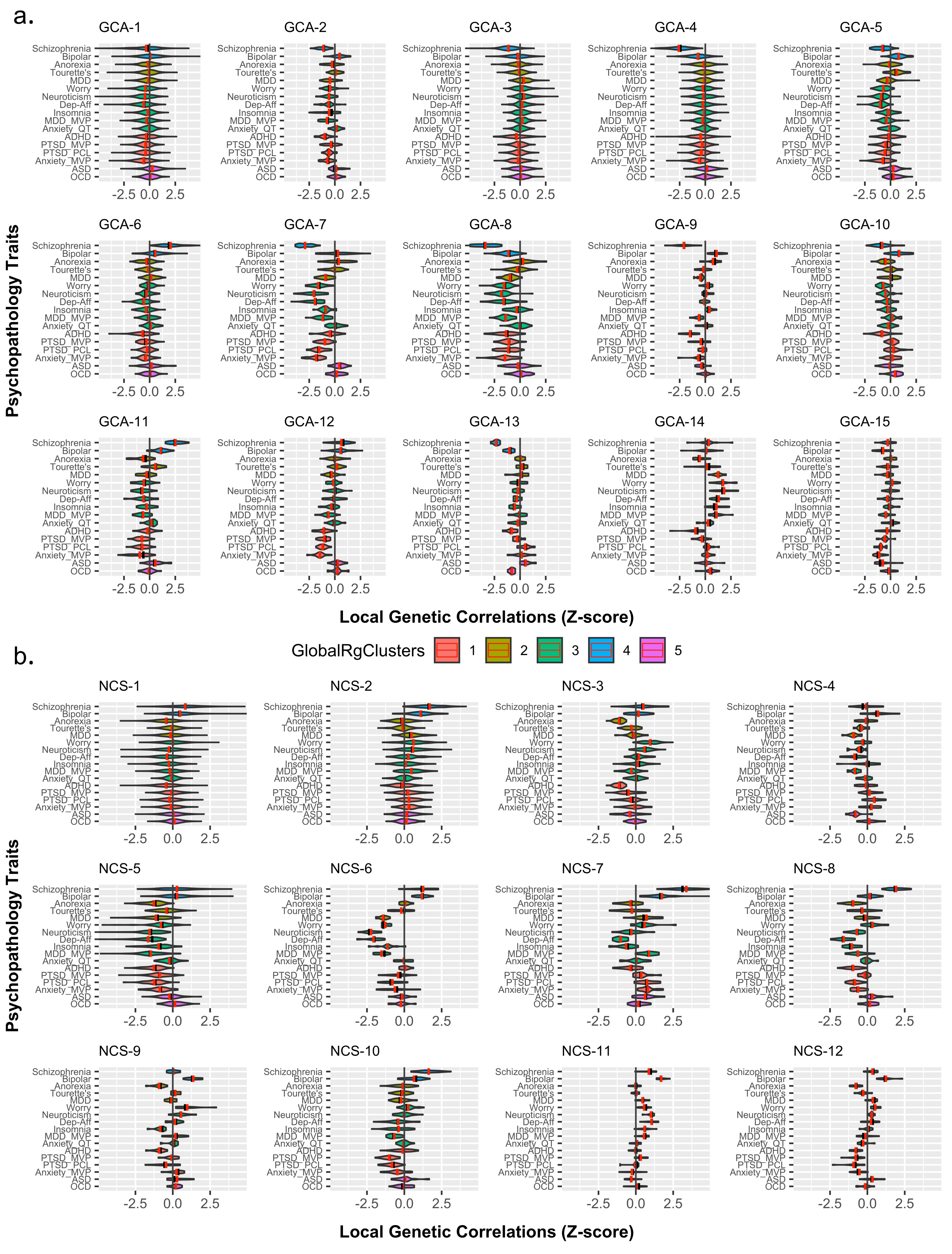

### FigS6

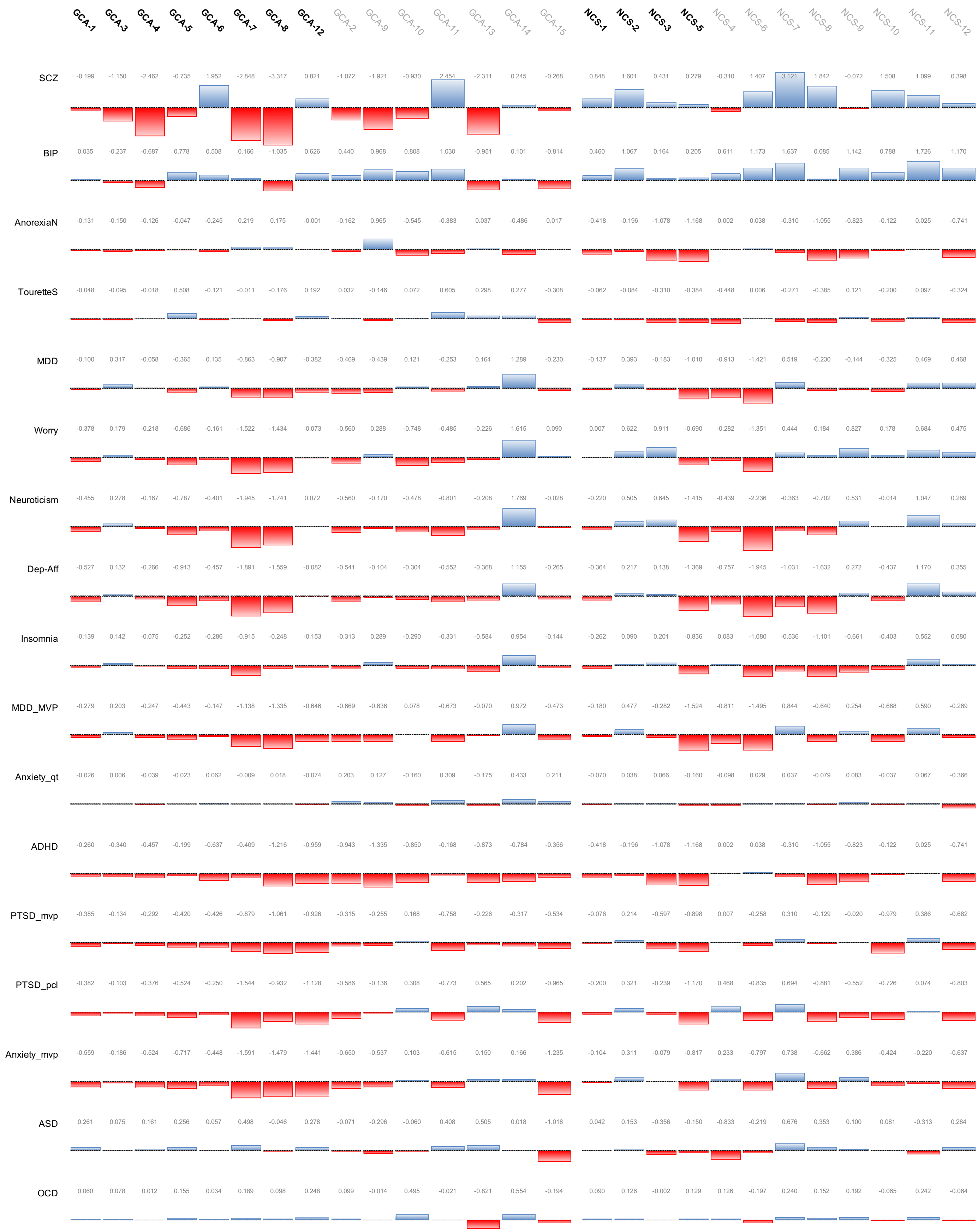
