## Supplementary Info for "Dissecting Biological Pathways of Psychopathology using Cognitive Genomics"

### Data Processing

##### *Data Curation*

To thoroughly examine the pleiotropic relationship between cognitive dimension and psychopathology, we consolidated 19 Cognitive Traits, 17 Psychopathological Traits, Education Attainment, and Socioeconomic Status GWAS summary statistics for the current study. Cognitive Traits included General Cognitive Ability[1] (indicated as MTAG_GCA in the report), ASSET Discordant[2], DeMange-Cognition[3] (shown as GSEM_GCA currently), and DeMange-Noncognition[3] (shown as GSEM_NCS currently). We also curated GWAS summary statistics for other cognitive traits. These include the GWAS summary statistics obtained via collaboration with Biogen Inc., Full-Scale IQ, Numeric Reasoning, Verbal Reasoning, General Cognitive Ability (Computed using approaches detailed in Davies et al., 2018), Pairs Matching, and Reaction Time. Data from Biogen Inc. were the most recent UK Biobank data freeze of cognitive tests. These similar cognitive traits were also available via our collaboration with the Institute of Behavior Genetics (IBG). Both sets of GWAS summary statistics were included for exploration because, for the latter, missing data were imputed for the entire UK Biobank data (Further details of the imputation method were reported in Hatoum et al. [4]). The following GWAS summary statistics were included from IBG, Executive Function, Digit Symbol, and Trail Making Test. Two sets of Education Attainment GWAS summary statistics were included; the first was reported by Lee et al. [5], and the second set was from UK Biobank's latest data freeze.  Finally, for follow-up and post-hoc investigation, GWAS summary statistics for the Townsend Deprivation Index were also included.

GWAS summary statistics psychopathological traits included Attention-Deficit/Hyperactivity Disorder[6], Anorexia Nervosa[7], Generalized Anxiety Disorder[8,9], Bipolar Disorder[10], Insomnia[11], Tourette's syndrome[12], Autism Spectrum Disorder[13], Major Depressive Disorder[14,15], Post-Traumatic Stress Disorder, and Schizophrenia[16]. Notably, two Mood disorder definitions were included. The first was reported by Howard et al. [14], and the second was obtained via collaboration with the Million Veterans Project[15]. The rationale for including both sets of Major Depressive Disorder GWAS was that the sample combinations were slightly different - the latter including data from the FinnGen study[15]. Similarly, for Generalized Anxiety Disorder, GWAS summary statistics from ANGST consortium[8] and the Million Veterans Project[9] were included. Over and above psychiatric traits, several personality dimensions (i.e., Neuroticism, Depressive-Affect, and Worry)[17] were added to accentuate the analysis.

##### *Summary Statistics Quality Control*

Because we obtained GWAS the summary statistics from various research groups and biobanks, we carried a series of data harmonization procedures for the GWAS summary statistics – ensuring that the data was aligned correctly for the analyses reported in the subsequent sections.
A summary statistics QC pipeline was written to perform the analysis and could be found at https://github.com/maxzylam/SumstatsQC. The procedures for SumstatsQC were loosely based on the EasyQC pipeline[18] and primarily written for use in a single virtual machine with multiple CPUs on Google Cloud. Dependencies for the pipeline were described on the GitHub README file.

To standardize summary statistics, the 1000 genome phase 3 reference panel was utilized. All variants on the phase 3 panels were assigned "CHR:BP:A1:A2" key ID for data merging in subsequent steps. After that, columns in the GWAS summary statistics were standardized to "UID SNP CHR BP A1 A2 FRQ INFO OR/BETA SE P Nca Nco". If the phenotypes were quantitative traits, a single N column was assigned automatically. Normalized Z-scores were estimated from the P-values of the GWAS summary statistics and assigned directions based on the Beta or Odds-Ratio. GWAS summary statistics were split into the 22 chromosomes to facilitate quicker processing time. To allow further standardization of the GWAS summary statistics, we excluded non-autosomal chromosomes. After Z-score estimation, SNP and allele definitions from the reference panel were merged with the GWAS summary statistics. This would allow us to check for strand flips or reference allele flips. All GWAS summary statistics were aligned against the reference alleles of the reference panel. Where necessary, effect estimates were flipped along with allele alignments by multiplying effect sizes by -1. An observation that we made for several sets of summary statistics processed in this manner was that the pipeline excluded variants with long indels alleles. The reason was that these indels were often truncated or represented differently from the 1000 genomes reference panel depending on how the original processing was carried out by the consortia or biobank in which the GWAS summary statistics were obtained.

In addition to allele alignments, where available, INFO score and allele frequencies were filtered from the GWAS summary statistics so that high-quality variants were retained in the subsequent analyses. We kept SNPs/variants with allele frequencies greater than 0.005 and INFO score greater than 0.3. In addition, we compared the allele frequency differences with the reference panel; variants with allele frequency greater than 0.15 were excluded. The procedures would increase our confidence that we were not interpreting results that might be population or sample-specific. In the case of ambiguous alleles, we excluded those that were above an allele frequency of 0.35. And finally, we excluded variants with allele frequencies that were 0.5.

The description of each of the GWAS summary statistics utilized in the current report is reported in **Supplementary Table 1: Summary of GWAS summary statistics.**

#### Data Reduction and Clustering Approaches for Cognitive and Psychopathological Traits

##### *Global Genetic Correlations (Cognitive Traits versus Psychopathology)*

19 Cognitive Traits and 17 Psychopathological Traits were included overall in the current study. To visualize the global genetic correlations, we carried out pairwise LD score regression with all traits. This was carried out via the genetic correlation matrix wrapper function that can be found in GenomicSEM::ldsc(), stand=TRUE function (GenomicSEM version 0.0.2, https://github.com/GenomicSEM/GenomicSEM). The wrapper function in GenomicSEM carries out pairwise global genetic correlations and represents the data in matrix form. The global genetic correlation matrix was reported in **Supplementary Table 2a: Global Genetic Correlations of Cognitive Traits and Psychopathology.**

Principal components and K-medoid partitioning cluster analyses were carried out on the genetic correlation matrix. PCA analysis revealed that the first two PCs captured 85.7% of the total variance. Loadings of each cognitive trait on the PCs were reported in **Supplementary Table 2b: K-Medoid Clustering and PCA Dimensions 1 and 2**.

##### *Cluster analyses for the global genetic correlation matrix*

Global genetic correlation analysis was first read into R as matrix *x* – where we proceeded to estimate the Euclidean distance matrix from. We selected two broad categories of clustering methodology that had been widely reported in academic literature (i) partitioning methods (ii) hierarchical clustering methods. Within partitioning clustering methodology, we estimated fit statistics for k-means and k-medoid clustering. And within hierarchical clustering methodology, we calculated fit statistics for agglomerative and divisive clustering. Initial clustering analyses were carried out via the FactoMineR and FactoExtra R packages (version 1.07.999, Le et al., 2008). We tested cluster solutions starting from $k = 2$ and increased the number of cluster solutions until one of the four methods yield a cluster with a single data point. The cluster prior to that was then designated as $k_{max}$. In this case, the maximum clusters were defined as $k_{max} = 5$. Fit statistics for each set of cluster analyses were estimated using the cqcluster.stats module from the fpc R package (version 2.2-9, Akhanli and Hennig[19]). The fit statistics used in the current report were extensively discussed by Akhanli and Hennig[9]. For straightforward interpretation of the fit statistics, we scaled the fit statistics such that larger metrics always meant better fit. All fit statistics were summed, based on previously established approaches[9], to get overall fit statistics for the cluster solutions. A five-cluster solution computed by k-means and k-medoids appeared to have comparable fit statistics. However, k-medoid cluster appeared to have slightly better stability after bootstrapping compared to the k-means solution. The cluster analysis results allowed further annotation of the genetic correlation matrix (See **Supplementary Figure 1**; **Supplementary Table 2c: Fit Statistics for Global RG clustering**).

#### GWAS-by-subtraction

We estimated the Non-Cognitive Skills latent factor based on the exact steps reported by Demange et al.[3] described at https://rpubs.com/MichelNivard/565885. The input GWAS summary statistics were, however, different. We used the publicly available Lee et al.[5] Education Attainment GWAS summary statistics (without the 23andMe data) and the most powered General Cognitive Ability GWAS (Lam et al. [1]). This was recommended by the original authors where new information should be added to aid in the definition of Non-Cognitive Skills. We also note that that the GWAS summary statistics that were included in GWAS-by-subtraction procedures were first quality controlled by earlier summary statistics QC procedures described above. As a sanity check to ensure that GWAS summary statistics were similar to those reported by Demange and colleagues[3], we carried out LD score regression[20] on the extracted latent factor scores compared to those that were reported on previously. The results indicate that the genetic correlation between the newly estimated summary statistics and those previously reported by Demange and colleagues[3] were the same ($R_g = 1$). Nevertheless, we noted that the power of Non-Cognitive Skills was slightly reduced. This could be related to two concurrent reasons. First, Demange and colleagues utilized data from Education Attainment that included the 23andMe data, which was sizable. Second, the GWAS summary statistics used earlier was a less powered version for General Cognitive Ability at 257K individuals, compared to that in the current report at 373k estimated sample size. Nonetheless, because the primary objectives of the present report were not loci discovery at the level of GWAS p-values, we made no further loci-based comparisons with the earlier study[3].

#### Local Genetic Correlation Analysis

Local genetic correlations were carried out via ρ-HESS, based on analytic steps described in https://huwenboshi.github.io/hess/local_rhog/. Local genetic correlations were computed on 2353 LD-independent regions across the genome. The LD independent regions were calculated via LD detect[21] using variants with minor allele frequencies greater than 0.05. Further details of the estimation of LD independent regions were previously reported[22]. Local genetic correlations were carried out with General Cognitive Ability and Non-Cognitive Skills with each of the 17 psychopathological traits selected for the current report (ρ-HESS version 0.5.4). For the purpose of the present report, we wrote a wrapper for ρ-HESS such that produces a series of helper scripts that allowed us to scale the analysis. The wrapper scripts could be found at https://github.com/maxzylam/rho-HESS-wrapper.

##### *ρ-HESS results*

Heritability of LD independent regions for General Cognitive Ability (GCA) and Non-Cognitive Skills (NCS) was reported in **Supplementary Table 3: Local heritability of General Cognitive Ability and Non-Cognitive Skills.** We noted that the summed heritability of each of the cognitive dimensions was consistent with those estimated by global genetic heritability. Summed heritability was calculated by taking the total heritability across all LD independent regions.

Local genetic correlations were represented in covariances and Z-scores by ρ-HESS. Due to the small values of the covariances, we opted to use the standardized Z scores for downstream analysis. The results of GCA and NCS were reported in **Supplementary Table 4: Local genetic correlations estimated by ρ-HESS for General Cognitive Ability and Psychopathology trait pairs** and **Supplementary Table 5. Local genetic correlations estimated by ρ-HESS for Non-Cognitive Skills and Psychopathology trait pairs.**

*Manhattan plots for ρ-HESS results*

There are two sets of Manhattan plots visualized for the local genetic correlation output (in Z-scores). The first had the x-axis aligned based on chromosome location and base-pair coordinates. In the second, results were sorted from lowest to highest local genetic prioritized meta-loci mentioned above (See **Supplementary Figure 2: Manhattan plots for ρ-HESS local genetic correlation output**).

In addition to Z-scores, we estimated the genetic correlations based on ρ-HESS outputs. Genetic correlations were estimated in the following manner

$$R_{g-HESS} = \frac{\Sigma(Cov_j)}{\Sigma(h_{ja}^2 * h_{jb}^2)}$$

Where $j$ is a given set of LD independent regions, $Cov$ represents the covariance at each LD independent region and, $h_j^2$ is the heritability of $j$ regions; $a$ represents cognitive dimensions, either GCA or NCS, and $b$ represents psychopathological trait.

##### *Local Genetic Correlations for the MHC region*

Due to the complex LD structure that was harbored within the MHC region, the region was excluded from subsequent cluster analysis. However, for completeness, the Manhattan plot of the MHC region was visualized. In **Supplementary Figure 3: Manhattan plots for ρ-HESS local genetic correlation output – MHC region**, we show plots that were aligned based on base-pair coordinates, as well as LD independent regions sorted based on the smallest and largest local genetic correlation Z-scores. There were 23 LD-independent regions that were part of the MHC region.

##### *Schizophrenia primarily related to GCA and NCS*

To further investigate local genetic correlations between cognitive dimensions (GCA/NCS) and psychopathological traits, we flagged LD-independent regions showing strong local genetic correlation Z-scores (|Z| > 4, correcting for multiple testing in 2353 LD independent regions, assuming Z = 1.96 represents p = 0.05, see **Supplementary Table 6: Local Genetic Correlations for General Cognitive Ability/Non-Cognitive skills and Psychopathological traits that have effect sizes |Z| > 4**). 88 LD-independent regions showed strong local genetic correlations between GCA and schizophrenia, 27 LD-independent regions showed strong local genetic correlations between NCS and schizophrenia. 13 LD-independent regions showed strong correlations with affective traits and GCA, and 27 LD-independent regions showed strong correlations with affective traits and NCS.

***Comparing global genetic correlations estimated by LDSC and ρ-HESS***

To examine if there was concordance between LDSC and ρ-HESS, we compared global genetic correlations estimated by both methods. Default parameters were carried out on LDSC, as part of standard procedures, summary statistics were pruned to 1.2 million HAPMAP3 SNPs for global genetic correlation estimations for LDSC. Bivariate genetic correlations were carried out for GCA and NCS versus each of the psychopathological traits included in the current study. To estimate global genetic correlation via ρ-HESS, we summed the estimated covariances across all LD-independent regions in the genome and the estimated heritability for either cognitive dimension (GCA/NCS) and psychopathological trait. A scatterplot was constructed to visualize the concordance between LDSC and ρ-HESS (**Supplementary Figure 2: Comparing global genetic correlations**)

***Heritability Benchmarks for GWAS summary statistics***

The heritability estimates for GCA and NCS summary statistics were compared with what is current reported in the literature, as well as across LDSC and ρ-HESS.

*General Cognitive Ability*

*Reported by Davies et al.* [23]*:*

The report included all common SNPs using GCTA-GREML in four of the largest individual samples: English Longitudinal Study of Ageing (ELSA: $N = 6661$, $h^2 = 0.12$, SE = 0.06), Understanding Society ($N = 7841$, $h^2 = 0.17$, SE = 0.04), UK Biobank Assessment Centre ($N = 86{,}010$, $h^2 = 0.25$, SE = 0.006), and Generation Scotland ($N = 6{,}507$, $h^2 = 0.20$, SE = 0.05)

*Reported by Savage et al.* [24]*:*

SNP heritability estimated for the entire sample $h^2_{SNP}$ was 0.19 (SE=0.01) estimated by LDSC.

*Estimation of Heritability within Current Study*

$h^2_{GCA\text{-}LDSC}$ = 0.1538 (0.0059)
$h^2_{GCA\text{-}HESS}$ = 0.2310 (1.11e-5)

*Non-Cognitive Skills*

*Reported by Demange et al.* [3]*:*

$\lambda_{NonCog\text{-}EA}$ = 0.2565 (Genomic SEM)

*Estimation of Heritability within Current Study:*

$\lambda_{NonCog\text{-}EA}$ = 0.230 (Genomic SEM)
$h^2_{NCS\text{-}LDSC}$ = 0.2035 (0.0105)

$h^2_{NCS\text{-}LDSC}$ = 0.310 (6.20e-5)

Both general cognitive ability and non-cognitive skills are still in and around the ballpark of what is reported in the literature. It is notable that ρ-HESS is estimating heritability slightly higher than LDSC. This appears consistent between GCA and NCS. The explanation for that is the additional 10% of heritability is likely related to ρ-HESS using genome-wide summary statistics rather than the 1.2 million HAPMAP3 SNPs that LDSC uses.

#### Density-Based Scan Procedures for Identification of Meta-Loci

Density-based Spatial Clustering of Applications with Noise (DBSCAN) procedures[25] was carried out for General Cognitive Ability and Non-Cognitive Skills separately. As previously indicated, the MHC region was excluded from these analyses. First, we performed data reduction on the local genetic correlations between the cognitive dimensions and psychopathological traits via Uniform Manifold Approximation and Projection for Dimension Reduction (UMAP, McInnes, et al., 2020). The method was implemented in R 3.6.3 via the uwot package (version 0.1.10). UMAP had previously been demonstrated to be superior in retaining data structure in comparison with other similar methods such as t-SNE (McInnes et al., 2020). We required the number of nearest neighbors to be five and estimated the minimum spread value for the 2353 LD-independent regions to be $mindist = \frac{1}{\sqrt{n\ regions}}$. We also assumed that approximately 70% of the manifolds are likely to show local connections. Data was reduced into two dimensions for GCA and NCS and appended to local genetic correlation results (See **Supplementary Table 4b: Local genetic correlations estimated by ρ-HESS for General Cognitive Ability and Psychopathology trait pairs** and **Supplementary Table 5b. Local genetic correlations estimated by ρ-HESS for Non-Cognitive Skills and Psychopathology trait pairs**). Thereafter, both UMAP dimensions were subjected to density-based scan procedures. As with earlier UMAP analysis, nearest neighbors were set to minimums of 5. To find the optimal set of meta-loci represented by the data, we carried out density-based scan procedures first for General Cognitive Ability. Epsilon (eps) radius for detecting clusters were varied in the procedure using a divisive clustering approach. Eps was set at maximums (eps = 3.5) such that only 1 cluster in the data set existed. Thereafter, the eps index was decreased in increments of 0.1, generating various meta-loci for GCA until we reached a point where the DBSCAN procedures generated *d* number of clusters with outliers. The optimal number of DBSCAN clusters was *d*-1, the optimal eps radius was defined as the metric just prior to outliers emerging in the DBSCAN clusters. DBSCAN clusters were defined as "meta-locus." The expectation was that DBSCAN procedures clustered LD-independent regions with distinct local genetic correlation profiles and localized by LD. To further examine if each LD-independent region was indeed clustered via local genetic correlation patterns, we visualized each meta-locus for GCA and NCS on genome-wide karyogram (See **Figure 2: UMAP and Karyogram Plots for Meta-Locus Definitions**). The visualization showed that regional distributions were well distributed across the genome and did not appear to be varying by LD or localized effects in the genome. These results increased our confidence that regional local genetic correlations were likely driving the DBSCAN clustering rather than LD patterning across the genome.

##### *Conceptual Description of the DBSCAN Clustering Algorithm*

An important distinction has to be made between conventional partitioning cluster analysis that was utilized for global genetic correlation analysis versus procedures applied to the local genetic correlation data is that we used DBSCAN for the local genetic correlation data. DBSCAN is a sequential clustering methodology that is more appropriate from highdimensional data that also tends to be fairly noisy. The DBSCAN algorithm following the following heuristic given a certain radius and minimum points per cluster parameter. For each data point, in this case, for each LD-independent region, DBSCAN estimates the distance relative to all 2353 LD-independent regions. If the distance is less than or equal to the epsilon, then the LD-independent region would be marked as a neighbor of x. If the LD-independent region gets a neighboring count greater than or equal to the minimum points per cluster, DBSCAN marks the region as a core point or visited. For each core point, if not already assigned to a cluster (meta-locus), then create a new cluster (meta-locus). DBSCAN then recursively finds all neighboring points and assign them to the same cluster (meta-locus) as the core point. These steps are iterated until all LD-independent regions were assigned cluster membership (or all points assigned to a meta-locus). For further details, see https://towardsdatascience.com/k-means-vs-dbscan-clustering-49f8e627de27, and http://www.sthda.com/english/wiki/wiki.php?id_contents=7940.

##### *Additional considerations regarding DBSCAN methodology*

By performing DBSCAN analysis using the approach that was reported above, the first meta-locus that was the largest for both GCA and NCS would include data points, or in this case, LD-independent regions that are weakly associated with other regions in terms of their local genetic correlation profiles. There could be two explanations for this. The first is that there could have been simply a power issue either for the psychopathological traits or either of the cognitive dimensions, where the local genetic correlation profile across traits is simply not strong enough to be separable into additional meta-loci. In this scenario, it would suggest that there are additional meta-loci awaiting further elucidation with more powered GWAS summary statistics in subsequent work. The second scenario would be that the first meta-locus is a very specific “large” meta-locus explaining specific biology. However, if we examine the percentage of heritability explained by either GCA or NCS, we will see that the first meta-locus harbors most of the heritability explained for either GCA or NCS. Along with the wide distribution of the local genetic correlation profile discussed in the preceding section, it would be fair to infer that the first scenario, which describes a situation of limited statistical power, is more likely. Larger sample sizes and more powered GWASs would be necessary to address the challenge.

##### *Visualization of Distribution of Local Genetic Correlations within Meta-Locus*

To further understand the nature of local genetic correlation distributions underlying each meta-locus, distributional patterns of the local genetic correlations with each set of LD-independent regions defined by the meta-loci in GCA or NCS were visualized (See **Supplementary Figure 5: Distributions of local genetic correlations for each LD-independent region within each meta-locus, Figure 3: Violin plots for Z-score distributions of Local Genetic Correlations within each prioritized meta-locus**). We displayed mean and median Z scores on the distributional patterns visualized in the violin plots. To examine which properties were separating the meta-loci, we computed the following distributional properties for each meta-locus displayed: Median, Median Absolute Deviation (MAD), Interquartile range, Min value, and Max value. We repeated PCA procedures described in Section 2.1. Instead of the global genetic correlation matrix, we entered the local genetic correlation Z scores matrix. We were then able to perform a regression analysis for each meta-locus specific PC and distributional properties of local genetic correlations. The results were generally unremarkable. One of the principal components would usually index the median local genetic correlation (median Z), and the second principal component would index an index of variance, either the Min or Max value, MAD or IQR. This is not unexpected given that no further information other than local genetic correlations were fed into UMAP and DBSCAN procedure. Z-scores were preferred when performing UMAP and DBSCAN procedures over local genetic correlations for each LD-independent region. The rationale was to cluster each LD-independent region based on the strength of the covariance of effect sizes. Estimating local genetic correlations would invariably involve weighting the covariance by the heritability of either GCA/NCS and that of 17 psychopathological traits. However, this would suggest that information regarding the association power for each region would be incorporated in the data reduction approach, which would potentially bias the clustering results. In the current study, we are leveraging the power of the cognitive dimensions as a means of interpreting results that might emerge from the local genetic correlation analysis and the downstream analyses. Nonetheless, within the prioritized meta-loci for GCA and NCS (Figure 3), we displayed local genetic correlations for each psychopathological trait within the meta-locus as a means of reference.

#### Functional Annotation and Gene Prioritization

Functional annotation and gene prioritization was carried out only for GWAS summary statistics indexing General Cognitive Ability and Non-Cognitive Skills. The rationale for carrying out gene prioritization in this manner was such that we could have a signal that is cognitive-centric rather than driven by factors that might be specific to or related to the psychopathological conditions investigated. Both GCA and NCS were estimated from the general population, hence, less likely to be influenced by extraneous clinical factors (e.g., illness trajectories and medication) that might have been uniquely driven by the psychopathological condition.

Gene prioritization approaches used considered standard in GWAS downstream analysis. These could be categorized into gene-based genome-wide association approaches (MAGMA and POPs), transcriptome-wide association approaches (S-PrediXcan, SMR/HEDI, and FOCUS transcriptome-based finemapping). In the current report, results of the downstream analysis were additionally assigned to the respective GCA or NCS meta-loci.

##### *MAGMA Gene-Based Genome-Wide Association (GBGWA)*

MAGMA gene-based genome-wide association analysis[27] was carried out for GCA and NCS. GWAS SNP-based summary statistics were used as input data for the MAGMA GBGWA analysis. Gene definitions based on b37 were utilized (see https://ctg.cncr.nl/software/magma). Note that for the current analysis, the latest version of MAGMA v1.08 was utilized. Further data analysis steps could be found on the provided website.

##### *Polygenic Priority Score (PoPs)*

PoPs is a gene prioritization method that leverages genome-wide signal from GWAS summary statistics and incorporates data from an extensive set of public bulk and single-cell expression datasets, curated biological pathways, and predicted protein-protein interactions. Methodological details of PoPs were previously described by Weeks and colleagues[28]. Data analytic steps are provided at (https://github.com/FinucaneLab/pops). PoPs leverages 57,543 gene features for prioritization. 40,546 features were derived from gene expression data, 8,718 features extracted from protein-protein interaction network, and 8,479 features based on pathway membership. PoPs was carried out for both GCA and NCS. Both MAGMA and PoPs results were reported in **Supplementary Table 8: MAGMA Gene-Based Genome-Wide Analysis and PoPs gene prioritization scores.**

##### *Summary Statistics Mendelian Randomization and Heterogeneity in Dependent Instruments Analysis*

SMR allows the indirect mediating effect of gene expression to be incorporated into the SNP/Variant phenotype effects, while HEIDI allows potential heterogeneity of the mediating effect caused by linkage to be also modeled in the analysis[29]. For the current report, we used eQTL annotations from the Brain e-META database (meta-analysis of GTEx, Common Mind Consortium, and ROSMAP brain eQTL data to be modeled; and the PsychENCODE data. Two versions of the PsychENCODE data were used HCP and PEER adjusted. The union of results from all three annotation databases was considered to minimize prioritization related to methodological variance (**Supplementary Table 9a-c: Summary Statistics Mendelian Randomization**). Note that because HEIDI assumes that genes with significant results were less likely to have eQTL mediating the SNP-phenotype effect due to linkage, we inverted the significant effects during the gene prioritization procedures.

***Summary Statistics PrediXcan (S-PrediXcan) Transcriptome Wide Analysis***

S-PrediXcan, (formerly known as MetaXcan) was carried out to leverage eQTL data for gene prioritization (Oct 16, 2020, version). Details of the S-PrediXcan methodology are now well established and can be found in the report by Barbeira and colleagues[30]. For the current analysis, we leveraged the latest GTEx8[31] eQTL database. However, to allow more focused functional annotations and gene prioritization processes, we only selected eQTL data for neural tissues. These include Anterior Cingulate Cortex, Amygdala, Caudate – Basal Ganglia, Cerebellum, Cerebellar Hemisphere, Cortex, Frontal Cortex, Hippocampus, Hypothalamus, Nucleus Accumbens, Putamen – Basal Ganglia, Spinal Cord, and Substantia Nigra (**Supplementary Table 10a-m: Results of S-PrediXcan transcriptome-wide analysis based on GTEx brain tissues**).

***FOCUS Transcriptome Finemapping Analysis***

The FOCUS[32] (Fine-mapping Of CaUsal gene Sets) transcriptome finemapping analysis was designed to identify credible genes based on eQTL annotations, leveraging state-of-art GWAS and transcriptomic based statistical finemapping approaches. For our analysis, we included all genes that were identified as credible genes as part of gene prioritization procedures. FOCUS finemapping procedures and eQTL annotations are available at https://github.com/bogdanlab/focus. To keep consistent with the eQTL annotations of other transcriptomic association methods that were used in the current report, we only selected finemapping results based on brain tissue expression of the Anterior Cingulate Cortex, Amygdala, Caudate – Basal Ganglia, Cerebellum, Cerebellar Hemisphere, Cortex, Frontal Cortex, Hippocampus, Hypothalamus, Nucleus Accumbens, Putamen – Basal Ganglia, Spinal Cord, and Substantia Nigra (see **Supplementary Tables 11a/b: FOCUS transcriptomic wide fine-mapping procedures**).

***Gene Ranking and Prioritization***

To rank and select pertinent genes for downstream analysis, we carried out a series of gene ranking procedures. This was achieved by taking the 50$^{th}$ percentile cutoff for MAGMA GBGWA, PoPs gene scores, transcriptome association methods, and taking the union of gene lists emerging from these methods with credible genes identified by FOCUS transcriptomic finemapping (**Supplementary Table 12a-c**). For transcriptomic association methods that utilized more than one eQTL annotation database for prioritization, we used the average gene rank across annotations. The 50$^{th}$ percentile cutoff and corresponding gene rank for MAGMA GBGWA was $8867_{GCA}/8870_{NCS}$, PoPs was $9164_{GCA}/9170_{NCS}$, and TWAS $3806_{GCA}/3771_{NCS}$ (see **Supplementary Table 12a: Gene ranks and corresponding percentile**). We then attempted to annotate genes that have been prioritized via the HUGO gene annotation data base[33], at this stage, each gene was assigned to their respective meta-locus based on their genomic coordinates (**Supplementary Table 12c: HUGO Gene**

**Annotations of top 50th percentile gene (union of all TWAS methods + FOCUS credible genes)**.

#### Gene Set Analyses

To further annotate putative biological mechanisms and processes underlying each meta-locus for either cognitive dimension (GCA/NCS), we carried out gene set analysis on gene lists assigned to each meta-locus. Three methods were used, GSEA (Gene Set Enrichment Analysis[34]), WebGestalt[35], and FUMA::GENE2FUNC[36]. Methods of these approaches are discussed in the Online Methodology section.

##### *Gene Set Enrichment Analysis (GSEA) procedures for gene set enrichment and identification of driver genes*

In the current section, further details regarding GSEA and PoPs gene scores are discussed. GSEA prioritizes genes via the following heuristic, given a defined gene list *L*, GSEA determines if the genes are randomly distributed throughout pathway *S* or primarily found at the distributional tails. This is achieved via estimating an enrichment score based on any given metric that represents correlation with a given phenotype. In the current study, the selected phenotypes were GCA and NCS. The gene-phenotype correlation was denoted in the current study via gene scores estimated by PoPs. Where the higher the metric, the more likely the gene was associated with GCA or NCS. However, it is necessary to note that PoPs gene score is unidirectional, unlike gene expression. For gene expression, strong effect sizes in either direction represent strong gene-phenotype associations. However, for PoPs gene score, negative scores do not denote stronger associations with the phenotype. Rather, negative scores denote poor gene-phenotype associations. Hence, for the PoPs gene score to be incorporated into the GSEA gene scoring algorithm, we carried inverse rank scoring to scale the PoPs gene score to represent the unidirectionality of the metric. Such that 0 represented poor gene-phenotype association and 1 represented the strongest gene-phenotype relationship (**Supplementary Table 13: Computation of inverse rank scores based on PoPs gene scores and 50th percentile selected TWAS genes**). Enrichment scores were obtained via GSEA, which uses a running sum statistic to estimate deviation from null. First, the gene list for each meta-locus would be ordered by PoPs gene score. GSEA then compares each gene within the meta-locus against each gene sequentially within pre-defined pathway genes. If a gene within a given meta-locus and pathway matches, the gene score is summed. If the gene is not represented in the pathway, the gene score is subtracted. The running hypothesis is that if a list of genes is random, the enrichment score would be likely to tend towards the null. Whereas if a list of genes is well represented within a given pathway, there would be a significant deviation from the null. The null enrichment score was estimated by randomly ordering the association metric with the gene list and re-computing the enrichment score. This was repeated 1000 times to get a null distribution. Significance testing was carried out by testing if a given enrichment score for a particular gene set significantly deviated from its null distribution. In addition to the enrichment score for each gene set, it was possible to identify “driver genes” via GSEA. Driver genes are the core of a gene set accounting for the enrichment signal. Driver genes could be identified as those whose running sum statistic deviates for a given gene set, farthest from the null.

Using GSEA as a strategy to identify driver genes for each gene set identified per meta-locus, coupled with the requirement for multiple gene set analysis to identify converging gene set, over and above earlier MAGMA GBGWA and transcriptomic methods, we were able to identify a very specific list of genes for each meta-locus that were putatively responsible for biological mechanisms that might be subserved within each GCA or NCS meta-locus. We restricted gene-set annotations to Gene Ontology gene sets (Cellular

Component, Molecular Function, and Biological Processes) available as part of the Molecular Signature Database version 7.2 annotations (**Supplementary Table 14a-g: Results of Gene Set Analysis)**.

#### BrainSpan Spatial-Temporal Gene Expression Analysis

As a function of earlier gene set analysis and gene prioritization approaches, we were able to identify that potentially GCA could have been associated with neurodevelopmental mechanisms, while NCS could have been associated with synaptic function. We tested the hypothesis, using driver genes identified by GSEA earlier, that the spatial-temporal gene expression of driver genes indexing neurodevelopmental mechanisms were likely to be significantly expressed prenatally, whereas driver genes that were responsible for the synaptic function would potentially be fairly stable across the lifespan, if not show a preponderance of expression in adulthood.

##### *BrainSpan data preparation*

BrainSpan data was access via https://www.brainspan.org/static/download.html. RNA-Seq Gencode v10 summarized to genes database, that include normalized gene expression was utilized for the analysis. For purposes of data analysis, we recoded the developmental stage into “Weeks” of development so that we could obtain a higher resolution of the spatial-temporal gene expression profile. All post-natal stages were converted to weeks using *Weeks = Years*52Weeks + 37 Gestational Weeks* which resulted in the *Weeks* variable ranging from 8 – 2117 weeks. Gene expression for driver genes within the prioritized meta-loci for GCA and NCS were extracted and aggregated by taking the mean expression of all driver genes within a given meta-locus. We were able to compute the mean expression for GCA and NCS overall and for each GCA and NCS prioritized meta-locus (see **Supplementary Figure 9: Brain Span validation analysis for each prioritized meta-locus**).

##### *Linear mixed modeling for evaluating longitudinal spatial-temporal gene expression trajectories*

To test for gene expression trends over the lifespan for GCA and NCS, and for each meta-locus, we carried out linear mixed modeling using the lme4[37] and lmeTest[38]. For the overall comparisons of General Cognitive Ability and Non-Cognitive Skills, we dummy coded genes falling into each respective category and the dummy coded variable as a *Trait* variable (GCA vs. NCS). Random effect estimator was denoted for individual subjects within the BrainSpan database. Because *Weeks* was a variable of interest, we did not covariate the analysis for the developmental stage. Sex was included as a covariate. The null model was specified as $Expression \sim \beta_1 Weeks + \beta_2 Sex + (1 \mid subject)$ and the alternative model to test for the effect of the *trait* was $Expression \sim \beta_1 Weeks * Trait + \beta_2 Weeks + \beta_3 Trait + \beta_4 Sex + (1 \mid subject)$. A significant interaction effect in the alternative model would suggest that different profiles of spatial-temporal gene expression across the lifespan were present between GCA and NCS, respectively. Post-hoc analysis was carried out to examine specific meta-locus within GCA and NCS that were driving the interaction effect. The model $Expression_{meta-locus} \sim \beta_1 Weeks + \beta_2 Sex + (1 \mid subject)$, where *meta-locus* represented each of the prioritized meta-locus, for GCA and NCS respectively was used to evaluate if gene-expression within the meta-locus was prenatal, adulthood, or lifetime. The corresponding expectation for each scenario would be a significant positive effect, significant negative effect, and not significant.

#### Chemoinformatic Annotation Analysis

To further characterize if driver genes within the meta-locus were "actionable" – i.e., having known evidence of utility where there are current pharmacological agents presently know to act on the proteins by the genes,  we annotated driver genes using annotations provided by Finan et al. [39]. The annotations included several levels of evidence for the identified driver gene being "druggable." Tier 1 genes incorporated the targets of approved drugs and drugs in clinical development, Tier 2 incorporated proteins closely related to drug targets or with associated drug-like compounds, Tier 3A tier incorporated extracellular proteins and members of key drug target families or genes that were in proximity (±50 kbp) to a known GWAS SNP and had an extracellular location, and Tier 3B incorporated extracellular proteins and members of key drug target families (**Supplementary Table 15a/b: Prioritized Meta Loci for General Cognitive Ability/Non-Cognitive Skills, Gene Sets, Driver Genes, and Gene Druggability Annotations from Finan et al 2017)**.

#### Literature Benchmarking

A dashboard that summarized results reported in the current study was created so the data could be visualized in an integrated manner (**Supplementary Table 16a: Consolidated Results for GCA and NCS Prioritized Meta-Loci**). The global and local genetic correlation results, as well as driver genes for each meta-locus, were first included. These represented the core findings of the report. We then accessed the GWAS catalogue[40] to examine if driver genes had been identified in any prior reports that were indexed in the database. That would allow us to determine if the genes might have already shown pleiotropic evidence across previous reports (**Supplementary Table 16b: GWAS Catalogue Annotations Accessed Jan 14, 2021**). We carried out a literature search on either univariate GWAS of each of the psychopathological traits examined in the current report, and recent studies have examined the pleiotropy between psychopathological traits. Univariate GWAS reports included ADHD[6], ASD[13], Schizophrenia[16], Bipolar Disorder[41], Insomnia[11], Neuroticism[17], Depressed Affect[17], Worry[17] and, MDD[15]. For studies examining pleiotropy, the PGC-Cross Disorder Group report[42], P-factor[43] and, the Genomic Atlas report, which investigated global pleiotropy[44], were selected for benchmarking with the current results. For the global pleiotropy report[44], we selected only cognitive or psychiatric-related traits. Gene sets or biological pathways identified in the previous reports were extracted and compiled to compare with those identified in the current study (**Supplementary Table 16c: Gene Sets and Pathways that have been identified by previous univariate or multivariate GWAS reports**). The gene sets and pathways were then manually examined, and further summarized by key descriptive terms, GATE*, DEV*, BEH*, SYNAP*, NERV*, NEURO*, AXO*, DEND*, TRANSP*.

### Supplementary Figures

**Supplementary Figure 1. Global Genetic Correlations and Dimension Reduction Procedures**
*Note:* Panel (a). Principal Components Analysis and loadings of cognitive traits on top two extracted PCs. Panel (b). Partitioned k-medoid cluster analysis for 18 psychopathological traits. Cluster 1: PTSD: Post-Traumatic Stress Disorder, Rexp: Re-Experiencing symptoms, MVP: Million Veteran Project, PCL: Total PCL Symptom Scores, Anxiety: Anxiety Disorder. Cluster 2: MDD: Major Depressive Disorder (Howard et al., 2019), Tourette's: Tourette's Syndrome, Anorexia: Anorexia Nervosa. Cluster 3: Dep-Aff: Depressive Affect, Anxiety_QT: Anxiety Disorder Symptom Factor Scores, MDD_MVP: Major Depressive Disorder (Million Veteran Project). Cluster 4: Bipolar: Bipolar Disorder. Panel (c). Global genetic correlation matrix re-ordered to reflect results of PCA and partition clustering, MTAG GCA and GSEM NCS were highlighted in red.

**Supplementary Figure 2. Comparing global genetic correlations**
*Noted:* Panels (a) and (b). x-axis genetic correlations estimated by LDSC; y-axis genetic correlations estimated by ρ-HESS . Intercept was set to 0 for both reference lines. anxiety_qt: Anxiety Symptom Factor Scores, anxiety_MVP: Anxiety Disorder from Million Veteran Project, asd: Autism Spectrum Disorder, ptsd_pcl: Post-Traumatic Stress Disorder - Total PCL scores, ptsd_mvp: Post-Traumatic Stress Disorder case control, both PTSD phenotypes were from the Million Veteran Project, adhd: Attention Deficit/Hyperactivity Disorder, mdd_mvp: Major Depressive Disorder from the Million Veteran Project, depaff: Depressive Affect, mdd: Major Depressive Disorder (Howard et al., 2019), scz: Schizophrenia (PGC3), tourette : Tourette's Syndrome, anorexia: Anorexia Nervosa, bipolar: Bipolar Disorder. Panel (b). Red highlighted points were anorexia (Anorexia Nervosa) and OCD.

**Supplementary Figure 3. Manhattan plots for ρ-HESS local genetic correlation output**
*Note:* Panel (a). Independent LD segments were aligned based on their chromosome and e base-pair positions sequentially. Panel (b). Local genetic correlation results and LD independent segments were aligned based on the lowest Z score to highest Z score. Panels (a) and (b): GCA – General Cognitive Ability (in red) and NCS – Non-Cognitive Skills (in turquoise). The y-axis of all traits except for schizophrenia was set to Z score between -5 to 5. Plots for schizophrenia have y-axis limits between -10 and 10. Anxiety Symptom: Anxiety Symptom Factor Scores, Anxiety MVP: Anxiety Disorder from Million Veteran Project, MDD_MVP: Major Depressive Disorder from the Million Veteran Project, MDD: Major Depressive Disorder (Howard et al., 2019),

**Supplementary Figure 4. Manhattan plots for ρ-HESS local genetic correlation output – MHC region**
*Note:* Panel (a). Independent LD segments were aligned based on their chromosome and base-pair positions sequentially. Panel (b). Local genetic correlation results and LD independent segments were aligned based on the lowest Z score to highest Z score. Panels (a) and (b): GCA – General Cognitive Ability (in red) and NCS – Non-Cognitive Skills (in turquoise). The y-axis of all traits except for schizophrenia was set to Z score between -5 to 5. Plots for schizophrenia have y-axis limits between -10 and 10. Anxiety Symptom: Anxiety Symptom Factor Scores, Anxiety MVP: Anxiety Disorder from Million Veteran Project, MDD_MVP: Major Depressive Disorder from the Million Veteran Project, MDD: Major Depressive Disorder (Howard et al., 2019),

**Supplementary Figure 5. Distributions of local genetic correlations within each meta-locus**
*Note:* Panels (a) and (b). GlobalRgClusters: k-medoid clusters generated from earlier partitioning cluster analysis conducted on the global genetic correlations with cognitive and psychopathological traits (See Supplementary Table 3, Figure 2). GCA: General Cognitive Ability, NCS: Non-Cognitive Skills, Bipolar: Bipolar Disorder, Anorexia: Anorexia Nervosa, Tourette's: Tourette's Syndrome, MDD: Major Depressive Disorder (Howard et al., 2019), Dep-Aff: Depressive Affect, MDD_MVP: Major Depressive Disorder (Million Veteran Project), Anxiety_QT: Anxiety Disorder (Symptom Factor Scores), PTSD_PCL: Post-Traumatic Stress Disorder (Total PCL scores – Million Veteran Project), PTSD_MVP: Post-Traumatic Stress Disorder (Million Veteran Project)

**Supplementary Figure 6. Median Z-scores of local genetic correlations within each meta-locus**
*Note:* Z-scores greater than 0 were colored blue, and Z-scores less than 0, were colored red. Columns greyed out were meta-loci not prioritized (See Supplementary Table 5b and 6b). Numbers above each bar denote Z-scores for the cognitive-psychopathology trait pair. GCA: General Cognitive Ability, NCS: Non-Cognitive Skills, SCZ: Schizophrenia, BIP: Bipolar Disorder, AnorexiaN: Anorexia Nervosa, TouretteS: Tourette's Syndrome, MDD: Major Depressive Disorder (Howard et al., 2018), Dep-Aff: Depressive Affect, MDD_MVP, Major Depressive Disorder (Million Veteran Project), Anxiety_qt: Anxiety Disorder (Symptom Factor Scores), PTSD_mvp: Post-Traumatic Stress Disorder (Million Veteran Project), PTSD_pcl: Post-Traumatic Stress Disorder (Million Veteran Project, Total PCL symptom scores), Anxiety_mvp: Anxiety Disorder (Million Veteran Project), ASD: Autism Spectrum Disorder, OCD: Obsessive Compulsive Disorder.

**Supplementary Figure 7. Local Genetic Correlations for SES**
*Note:* Meta-loci 1,3,4,5,6,7,8,12 was prioritized for General Cognitive Ability, and Meta-loci 1,2,3,5 was prioritized for Non-Cognitive Skills based on their heritability (See Supplementary Table 5b and 6b).

**Supplementary Figure 8. Meta-Locus definition for represented as a function of local genetic correlations**
*Note:* Prioritized General Cognitive Ability Meta-Loci represented in peach color, and Non-Cognitive Skills Meta-Loci represented in blue. Meta-Loci definitions were represented based on binned local genetic correlations for each meta-locus. Size of font in each cell represented the strength of local genetic correlations, red fonts denoted local genetic correlations going in the negative direction and blue fonts denoted local genetic correlations going in the positive direction. GCA: General Cognitive Ability, NCS: Non-Cognitive Skills, Bipolar: Bipolar Disorder, Anorexia: Anorexia Nervosa, Tourette's: Tourette's Syndrome, MDD: Major Depressive Disorder (Howard et al., 2019), Dep-Aff: Depressive Affect, MDD_MVP: Major Depressive Disorder (Million Veteran Project), Anxiety_QT: Anxiety Disorder (Symptom Factor Scores), PTSD_MVP: Post-Traumatic Stress Disorder (Million Veteran Project), PTSD_PCL: Post-Traumatic Stress Disoder (Million Veteran Project, Total PCL symptom scores), Anxiety_MVP: Anxiety Disorder (Million Veteran Project), ASD: Autism Spectrum Disorder, OCD: Obsessive Compulsive Disorder.

**Supplementary Figure 9. Brain Span validation analysis for each prioritized meta-locus.**
*Note:* Panels (a), (b), (c). y-axis: normalized gene expression, x-axis: developmental timing in weeks. Panel (c). Output of Linear mixed model analysis for expression of overall General Cognitive Ability and Non-Cognitive Skills driver genes.

#### Supplementary Tables

**Supplementary Table 1.**
Summary of GWAS Summary Statistics

**Supplementary Table 2a.**
Global Genetic Correlations of Cognitive Traits and Psychopathology
**Supplementary Table 2b.**
K-Mediod Clustering PCA Dimensions 1 and 2
**Supplementary Table 2c.**
Fit Statistics for Global RG clustering

**Supplementary Table 3.**
Local heritability of General Cognitive Ability and Non-Cognitive Skills

**Supplementary Table 4a.**
Descriptive Statistics for GCA meta-loci
**Supplementary Table 4b.**
Local genetic correlations estimated by HESS for 17 General Cognitive Ability - Psychopathology trait pairs

**Supplementary Table 5a.**
Descriptive Statistics for NCS meta-loci
**Supplementary Table 5b.**
Local genetic correlations estimated by HESS for 17 General Cognitive Ability - Psychopathology trait pairs

**Supplementary Table 6**
Local Genetic Correlations for General Cognitive Ability/Non-Cognitive skills and Psychopathological traits that have effect sizes |Z| > 4

**Supplementary Table 7**
Local Genetic Correlations for General Cognitive Ability and Non-Cognitive Skills with Social Deprivation

**Supplementary Table 8**
MAGMA Gene Based Genome Wide Analysis and PoPs gene prioritization scores

**Supplementary Table 9a**
Summary Statistics Mendelian Randomization (Annotation: Brain e-Meta)
**Supplementary Table 9b**
Summary Statistics Mendelian Randomization (Annotation: PsychEncodeHCP)
**Supplementary Table 9c**
Summary Statistics Mendelian Randomization (Annotation: PsychEncodePEER)

**Supplementary Table 10a**
Results of S-PrediXcan transcriptome wide analysis based on GTEx brain tissues: Anterior Cingulate Cortex
**Supplementary Table 10b**
Results of S-PrediXcan transcriptome wide analysis based on GTEx brain tissues: Amygdala
**Supplementary Table 10c**
Results of S-PrediXcan transcriptome wide analysis based on GTEx brain tissues: Caudate – Basal Ganglia

**Supplementary Table 10d**
Results of S-PrediXcan transcriptome wide analysis based on GTEx brain tissues: Cerebellum
**Supplementary Table 10e**
Results of S-PrediXcan transcriptome wide analysis based on GTEx brain tissues: Cerebellar Hemisphere
**Supplementary Table 10f**
Results of S-PrediXcan transcriptome wide analysis based on GTEx brain tissues: Cortex
**Supplementary Table 10g**
Results of S-PrediXcan transcriptome wide analysis based on GTEx brain tissues: Frontal Cortex
**Supplementary Table 10h**
Results of S-PrediXcan transcriptome wide analysis based on GTEx brain tissues: Hippocampus
**Supplementary Table 10i**
Results of S-PrediXcan transcriptome wide analysis based on GTEx brain tissues: Hypothalamus
**Supplementary Table 10j**
Results of S-PrediXcan transcriptome wide analysis based on GTEx brain tissues: Nucleus Accumbens
**Supplementary Table 10k**
Results of S-PrediXcan transcriptome wide analysis based on GTEx brain tissues: Putamen – Basal Ganglia
**Supplementary Table 10l**
Results of S-PrediXcan transcriptome wide analysis based on GTEx brain tissues: Spinal Cord
**Supplementary Table 10m**
Results of S-PrediXcan transcriptome wide analysis based on GTEx brain tissues: Substantia Nigra

**Supplementary Table 11a**
FOCUS transcriptomic wide fine-mapping procedures
Overall results for General Cognitive Ability
**Supplementary Table 11b**
FOCUS transcriptomic wide fine-mapping procedures
Overall results for Non-Cognitive Skills

**Supplementary Table 12a**
Gene ranks and corresponding percentile
**Supplementary Table 12b**
Gene ranks based on multiple transcriptomic wide analysis approaches on General Cognitive Ability and Non-Cognitive skills GWAS Summary Statistics
**Supplementary Table 12c**
HUGO Gene Annotations of top 50th percentile gene (union of all TWAS methods + FOCUS credible genes

**Supplementary Table 13**
Computation of inverse rank scores based on PoPs gene scores and 50th percentile selected TWAS genes

**Supplementary Table 14a**
Candidate gene sets identified by GSEA, WebGestalt, and FUMA Gene2Func
**Supplementary Table 14b**
GO Biological Process Gene Set Annotations from GSEA, WebGestalt, and FUMA GENE2FUNC for General Cognitive Ability genes
**Supplementary Table 14c**
GO Molecular Function Gene Set Annotations from GSEA, WebGestalt, and FUMA GENE2FUNC for General Cognitive Ability genes
**Supplementary Table 14d**
GO Cellular Component Gene Set Annotations from GSEA, WebGestalt, and FUMA GENE2FUNC for General Cognitive Ability genes
**Supplementary Table 14e**
GO Biological Process Gene Set Annotations from GSEA, WebGestalt, and FUMA GENE2FUNC for Non-Cognitive Skills genes
**Supplementary Table 14f**
GO Molecular Function Gene Set Annotations from GSEA, WebGestalt, and FUMA GENE2FUNC for Non-Cognitive Skills genes
**Supplementary Table 14g**
GO Cellular Component Gene Set Annotations from GSEA, WebGestalt, and FUMA GENE2FUNC for Non-Cognitive Skills genes

**Supplementary Table 15a**
Prioritized Meta Loci for General Cognitive Ability, Gene Sets, Driver Genes, and Gene Druggability Annotations from Finan et al 2017
**Supplementary Table 15b**
Prioritized Meta Loci for Non-Cognitive Skills, Gene Sets, Driver Genes, and Gene Druggability Annotations from Finan et al 2017

**Supplementary Table 16a**
Consolidated Results for GCA and NCS Prioritized Meta-Loci
**Supplementary Table 16b**
GWAS Catalogue Annotations Accessed Jan 14, 2021.
**Supplementary Table 16c**
Gene Sets and Pathways that have been identified by previous univariate or multivariate GWAS reports
